# Social Determinants of Health in HIV/HBV Coinfection Compared with HIV and HBV Monoinfection: A Framework for Dynamic Individual-Level Social Vulnerability

**DOI:** 10.64898/2026.08.31.26361856

**Authors:** George A. Yendewa, Tayoot Chengsupanimit, Ali Dehghani, Ali Ahmed, Amir M. Mohareb, Michael L. Freeman, Chari Cohen, Ighovwerha Ofotokun, Karine Dubé

## Abstract

Human immunodeficiency virus (HIV) and hepatitis B virus (HBV) coinfection is associated with accelerated liver disease, but whether coinfection is associated with newly documented social determinants of health (SDoH) is unclear. We conducted a retrospective cohort study using TriNetX across 110 U.S. healthcare organizations (2010-2026). We propensity score matched adults with HIV/HBV to adults with HIV or HBV monoinfection. We organized newly documented SDoH indicators using a dynamic individual-level framework with four clinically recognized domains of social disadvantage: material vulnerability, healthcare access and engagement, interpersonal adversity, and psychosocial vulnerability. Matched cohorts included 10,071 HIV/HBV-HIV pairs and 9,659 HIV/HBV-HBV pairs (mean age, 47 years; 79% male; 66% non-White; median follow-up, 3.3 years). Over 178,900 person-years, HIV/HBV was associated with higher risk of the primary SDoH composite compared with HIV (11.5% vs 9.7%; incidence rate, 2.50 vs 1.97 per 100 person-years; hazard ratio [HR], 1.25; 95% confidence interval [CI], 1.15-1.37) and HBV (11.0% vs 6.4%; incidence rate, 2.39 vs 1.67; HR, 1.50; 95% CI, 1.35-1.67). HIV/HBV was also associated with higher material vulnerability and healthcare access and engagement composites in both comparisons, including housing instability, food insecurity, financial insecurity, insurance instability, and care disengagement/nonadherence (HR range, 1.22-3.33 vs HIV; 1.31-1.94 vs HBV). In the HBV comparison, HIV/HBV was additionally associated with interpersonal adversity, primary support stressors, and violence or victimization (HR range, 1.36-2.16). Findings were robust across sensitivity analyses. HIV/HBV was associated with more newly documented SDoH than monoinfection, supporting dynamic SDoH assessment.

## INTRODUCTION

Human immunodeficiency virus (HIV) and hepatitis B virus (HBV) frequently co-occur due to shared transmission pathways and structural determinants of exposure and care [1,2]. Globally, approximately 2.7 million people live with both infections, including 5% to 15% of people with HIV (PWH) in the United States [1–3]. HIV/HBV coinfection is associated with worse liver-related outcomes, including cirrhosis, hepatocellular carcinoma, and liver-related mortality, and requires complex long-term care [3–5]. This care burden is increasingly shaped by social determinants of health (SDoH). Among PWH, material insecurity, access barriers, and care disengagement are associated with poor adherence and lower viral suppression [6,7], while interpersonal adversities, including adverse childhood experiences, intimate partner violence, and disrupted social support, may further undermine long-term self-management [8,9]. Among people with HBV, structural barriers contribute to delayed linkage to care, suboptimal hepatocellular carcinoma surveillance, and low treatment uptake [10–12]. These concerns are especially relevant in HIV/HBV coinfection, where sustained engagement is required to maintain dual viral suppression and monitor liver disease progression [5,13]. Whether this clinical complexity is accompanied by an increased burden of newly documented social vulnerability over time remains unclear.

This uncertainty is partly methodological. Social risk has historically been poorly captured in administrative and electronic health record (EHR) data [14,15]. Earlier International Classification of Diseases, Ninth Revision (ICD-9) V codes offered limited granularity, and documentation was sparse and inconsistent [14,15]. The transition to ICD-10-CM Z codes substantially expanded the taxonomy, enabling structured documentation of social adversity [15,16]. Despite this, SDoH documentation remains markedly underused, appearing in only approximately 1% to 3% of medical records, and is often used to signal clinical complexity rather than systematic social risk screening [16,17]. In HIV, HBV, and HIV/HBV care settings, this underascertainment may be especially consequential, as social vulnerabilities that disrupt antiviral treatment adherence, hepatocellular carcinoma surveillance, and retention in care may remain undocumented until they emerge as downstream clinical risk [5,13].

Several converging lines of evidence suggest that HIV/HBV coinfection may generate, intensify, or reveal social vulnerability during longitudinal care, rather than merely co-occurring with pre-existing social disadvantage. Managing two chronic infections, including lifelong HBV-active antiretroviral therapy, hepatocellular carcinoma surveillance, and stigma related to both diagnoses may exacerbate existing financial strain, disrupt care engagement, and increase social instability [3–5,18,19]. Chronic illness independently increases housing and food insecurity, and multimorbidity produces cycles of income loss and care rationing that erode financial reserve [18,19]. In people with HIV/HBV coinfection, these demands may be compounded by layered HIV and HBV stigma, which have been associated with social withdrawal, disclosure burden, and care avoidance [20–22]. The syndemic framework recognizes that co-occurring diseases interact with social conditions to produce mutually reinforcing adverse outcomes [23]. Prior HIV/HBV studies also suggest that socioeconomic and behavioral factors independently predict liver disease progression [13,24]. However, comparative data examining SDoH in HIV/HBV versus HIV or HBV monoinfection remain limited.

Equally important is how social vulnerability is conceptualized. Current approaches mostly rely on static, area-level indices that aggregate census-derived characteristics to rank geographic disadvantage [25–28]. Although these measures predict population-level risk, they may miss individual-level social needs that emerge during chronic disease care; in prior work, sensitivity for detecting patient-reported social risks rarely exceeded 42% [28]. This distinction is especially relevant in HIV/HBV coinfection, where lifelong antiretroviral therapy, ongoing HIV and HBV monitoring, hepatocellular carcinoma surveillance, specialty care, layered HIV/HBV stigma, and cumulative treatment, time, cost, and quality-of-life burdens may create conditions through which social vulnerability emerges over time [3–5,20–22,31]. These burdens may precipitate or worsen material vulnerability, healthcare access disruption, interpersonal adversity, and social instability [6,7,18,19,29,30]. In this context, food insecurity, housing instability, insurance disruption, and care disengagement may be time-varying, recurrent, variable in intensity, clustered across domains, and potentially actionable through case management and social services integration [29,30]. A framework that captures these dynamic attributes may better identify when social vulnerability emerges, how it accumulates across domains, and where intervention could reduce morbidity in people requiring sustained longitudinal care.

In this study, we used large-scale real-world cohort data and a prespecified dynamic four-domain individual-level conceptual framework to compare documented SDoH outcomes among adults with HIV/HBV coinfection versus HIV and HBV monoinfection in the United States. We hypothesized that HIV/HBV coinfection would be associated with a higher incidence of documented SDoH indicators compared with either HIV or HBV monoinfection, reflecting the compounded burden of dual infection within a social context already shaped by structural disadvantage.

## METHODS

### Cohort Definitions

We included adults aged ≥18 years with qualifying events recorded on or after January 1, 2010. To ensure sufficient baseline observation and outcome ascertainment, eligible individuals were required to have ≥6 months of healthcare activity in the EHR. We defined HIV using ICD-10-CM codes for HIV infection. We defined chronic HBV using ICD-10-CM codes for chronic hepatitis B infection and/or laboratory evidence of persistent infection, defined as positive hepatitis B surface antigen or detectable HBV DNA ≥6 months. We defined three analytic cohorts: (1) HIV/HBV coinfection, requiring concurrent evidence of both HIV and chronic HBV; (2) HIV monoinfection, requiring HIV without any HBV diagnosis codes or positive HBV laboratory results; and (3) HBV monoinfection, requiring chronic HBV without any evidence of HIV. We excluded individuals with hepatitis C virus infection or prior liver transplantation. The HIV/HBV definition was applied consistently across both comparisons; comparator cohorts were defined separately. Complete code lists of cohort definitions are provided in the Supplementary File.

### Conceptual Framework and Outcome Domains

Our conceptual framework (Figure 1) was informed by the World Health Organization Commission on SDoH [25], the Healthy People 2030 SDoH framework [26], and syndemic theory [23]. We conceptualized social vulnerability as a dynamic, individual-level construct rather than a fixed area-level attribute. We defined dynamic social vulnerability as SDoH that may arise after cohort entry, vary over time, recur, differ in intensity, cluster across domains, and remain potentially modifiable through clinical or social-service intervention. Within this framework, documentation was treated as a clinically recognized marker of emerging vulnerability during HIV/HBV care.

**Figure 1.**
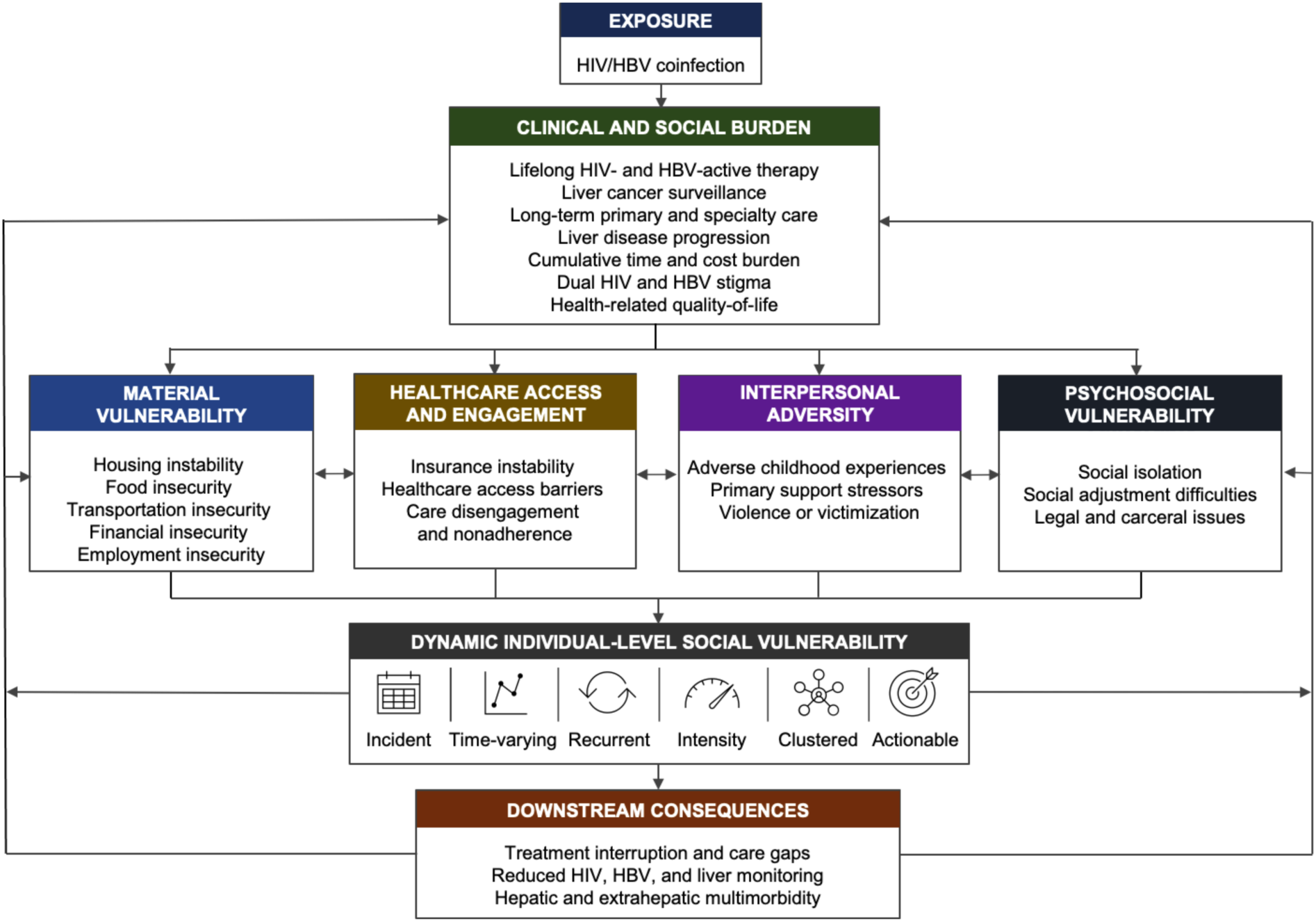
Conceptual Framework for Dynamic Individual-Level Social Vulnerability in HIV/HBV Coinfection. HIV/HBV coinfection is conceptualized as an exposure that may generate clinical and social burden through lifelong HIV- and HBV-active therapy, HCC surveillance, long-term primary and specialty care, dual HIV and HBV stigma, disease progression, and health-related quality-of-life burden. These burdens may contribute to four interrelated domains of social vulnerability: material vulnerability, healthcare access and engagement, interpersonal adversity, and social and environmental vulnerability. Collectively, these domains define a dynamic individual-level social vulnerability phenotype characterized by incident onset, time variation, recurrence, intensity, clustering, and clinical actionability. Potential downstream consequences include treatment interruption and care disengagement, reduced viral and liver disease monitoring continuity, and hepatic and extrahepatic multimorbidity.

The primary outcome was newly documented social vulnerability composite, defined as the first documentation of any qualifying SDoH indicator after the index date. Secondary outcomes included four domain-specific composites and their individual component outcomes. Domain-specific composites were defined as the first documentation of any component outcome within that domain after index. We hypothesized that social vulnerability in chronic viral coinfection arises through partially overlapping pathways across four prespecified domains. We organized these domains and their individual components as follows: material vulnerability, healthcare access and engagement, interpersonal adversity, and psychosocial vulnerability. Complete code lists for all outcomes are provided in the Supplementary File.

Material vulnerability was defined as unmet basic resource needs that may constrain stable living conditions, nutrition, transportation, employment, and the ability to sustain longitudinal HIV/HBV care [6,7,18,29,30]. We operationalized its individual components as housing instability, food insecurity, transportation insecurity, financial insecurity, and employment insecurity. The healthcare access and engagement domain captured barriers to obtaining, financing, and remaining engaged in medical care. Its individual components were insurance instability, healthcare access barriers, and care disengagement or nonadherence. Interpersonal adversity was defined as relational or family-level stressors that may affect disclosure, support, safety, and self-management. Its individual components were adverse childhood experiences, primary support stressors, and violence or victimization. Lastly, the psychosocial vulnerability domain encompassed broader social-contextual problems distinct from direct interpersonal relationships. Its individual components were social isolation, social adjustment difficulties, and legal or carceral issues. This domain was intended to reflect disruptions in social integration, community participation, adaptation to social environments, and legal or carceral circumstances that may affect care continuity and monitoring access [6,7,32]. Individual component outcomes of each domain were analyzed separately.

### Covariates and Propensity Score Matching

We performed 1:1 nearest-neighbor propensity score matching without replacement for each comparison to reduce baseline differences between HIV/HBV and comparator cohorts and to account for factors that could influence both coinfection status and subsequent SDoH documentation. For the primary analyses, we selected covariates *a priori* and grouped them into four categories: (1) demographics (age, sex, race, ethnicity); (2) medical comorbidities (liver disease, diabetes, hypertension, heart failure, ischemic heart disease, cerebrovascular disease, chronic lower respiratory disease, chronic kidney disease, neoplasms); (3) mental health diagnoses (anxiety disorders, major depressive disorder, post-traumatic stress and trauma-related disorders, bipolar disorder, schizophrenia and psychotic disorders); and (4) substance use conditions (nicotine dependence, alcohol, opioid, cocaine, cannabis, other psychoactive substance, and stimulant-related disorders). We included mental health diagnoses and lifestyle-associated risk factors given their known associations with both SDoH exposure and healthcare utilization, and the potential for confounding if individuals with greater psychosocial complexity were more likely to have both coinfection and SDoH documentation. In sensitivity analyses, we evaluated expanded covariate specifications incorporating virologic, treatment, and healthcare utilization measures. We assessed covariate balance using standardized mean differences, with values < 0.10 considered adequate balance.

### Sensitivity Analyses

We conducted three prespecified sensitivity analyses to evaluate robustness. First, we performed a 6-month landmark analysis, with outcome ascertainment beginning 6 months after index, to reduce misclassification from social vulnerabilities present before cohort entry but first documented during early evaluation. Second, we repeated analyses with additional adjustment for virologic and treatment variables, including CD4 count, HIV RNA, HBV DNA, antiretroviral therapy, and HBV-active antiviral therapy, to account for baseline disease severity, viral activity, and treatment exposure. Third, because greater healthcare use may create more opportunities for SDoH documentation, we performed a healthcare utilization-adjusted analysis accounting for baseline outpatient, emergency department, inpatient, and laboratory encounters before index.

### Statistical Analysis

We conducted all analyses using the TriNetX Advanced Analytics platform. We summarized continuous variables as mean ± standard deviation or median (interquartile range) and categorical variables as frequencies and percentages. We calculated incidence rates (IRs) per 100 person-years with corresponding 95% confidence intervals (CIs). For the primary analyses, we estimated time-to-event associations using Cox proportional hazards regression models and reported hazard ratios (HRs) with 95% CIs. We evaluated the proportional hazards assumption using Schoenfeld residuals. Two-sided p-values < 0.05 were considered significant.

## RESULTS

### Baseline Characteristics of HIV/HBV and Comparator Cohorts

For the HIV comparison, we identified 10,080 adults with HIV/HBV coinfection and 279,283 adults with HIV monoinfection before matching (Supplementary Table S1). In the unmatched cohorts, individuals with HIV/HBV differed from those with HIV monoinfection across demographic, clinical, mental health, and lifestyle-associated characteristics. After propensity score matching, 10,071 individuals with HIV/HBV were matched to 10,071 individuals with HIV monoinfection (Table 1). The matched cohorts were well balanced, with similar age at index, sex distribution, race and ethnicity, comorbidities, mental health diagnoses, and substance use-related conditions. For the HBV comparison, before matching, we identified 9,663 adults with HIV/HBV coinfection and 426,955 adults with HBV monoinfection (Supplementary Table S2). After propensity score matching, 9,659 individuals with HIV/HBV were matched to 9,659 individuals with HBV monoinfection (Table 1). Baseline characteristics were balanced after matching, with standardized differences < 0.1. In both the HIV and HBV comparisons, mean age was 47 years, 79% were male, and approximately 66% were non-White.

**Table 1.** Baseline Characteristics of Matched Adults With HIV/HBV Coinfection Compared With HIV and HBV Monoinfection.

| Variables | HIV/HBV vs HIV |  |  |  | HIV/HBV vs HBV |  |  |  |
| --- | --- | --- | --- | --- | --- | --- | --- | --- |
|  | HIV/HBV | HIV | p-Value | Standard difference | HIV/HBV | HBV | p-Value | Standard difference |
| **Totals** | 10,071 | 10,071 |  |  | 9,659 | 9,659 |  |  |
| **Age at Index (years)** | 47.1 ± 12.9 | 47.3 ± 13.8 | 0.246 | 0.016 | 47.0 ± 12.9 | 47.4 ± 14.3 | 0.074 | 0.026 |
| **Gender** |  |  |  |  |  |  |  |  |
| Male | 7,984 (79.3%) | 7,935 (78.8%) | 0.080 | 0.012 | 7,639 (79.1%) | 7,680 (79.5%) | 0.080 | 0.010 |
| Female | 2,082 (20.7%) | 2,151 (21.4%) | 0.230 | 0.018 | 2,015 (20.9%) | 1,979 (20.5%) | 0.367 | 0.018 |
| **Race or ethnicity** |  |  |  |  |  |  |  |  |
| White | 3,368 (33.4%) | 3,488 (34.6%) | 0.074 | 0.025 | 3,281 (34.0%) | 3,345 (34.6%) | 0.332 | 0.014 |
| Asian | 417 (4.1%) | 421 (4.2%) | 0.888 | 0.002 | 410 (4.2%) | 408 (4.2%) | 0.943 | 0.001 |
| Unknown Race | 1,596 (15.8%) | 1,628 (16.2%) | 0.539 | 0.009 | 1,574 (16.3%) | 1,563 (16.2%) | 0.830 | 0.003 |
| Black or African American | 4,337 (43.1%) | 4,230 (42.0%) | 0.120 | 0.022 | 4,069 (42.1%) | 3,998 (41.4%) | 0.300 | 0.015 |
| Hispanic or Latino | 477 (4.7%) | 473 (4.7%) | 0.894 | 0.002 | 467 (4.8%) | 500 (5.2%) | 0.276 | 0.016 |
| **Comorbidities** |  |  |  |  |  |  |  |  |
| Hypertensive diseases | 2,237 (22.2%) | 2,320 (23.0%) | 0.091 | 0.038 | 2,140 (22.2%) | 2,181 (22.6%) | 0.479 | 0.010 |
| Ischemic heart diseases | 562 (5.6%) | 564 (5.6%) | 0.951 | 0.001 | 540 (5.6%) | 538 (5.6%) | 0.950 | 0.001 |
| Heart failure | 308 (3.1%) | 292 (2.9%) | 0.507 | 0.009 | 293 (3.0%) | 299 (3.1%) | 0.802 | 0.004 |
| Diabetes mellitus | 864 (8.6%) | 919 (9.1%) | 0.172 | 0.019 | 844 (8.7%) | 867 (9.0%) | 0.560 | 0.008 |
| Chronic lower respiratory diseases | 1,054 (10.5%) | 1,101 (10.9%) | 0.284 | 0.015 | 1,007 (10.4%) | 935 (9.7%) | 0.085 | 0.025 |
| Chronic kidney disease | 736 (7.3%) | 705 (7.0%) | 0.397 | 0.012 | 711 (7.4%) | 687 (7.1%) | 0.505 | 0.010 |
| Diseases of liver | 790 (7.8%) | 803 (8.0%) | 0.734 | 0.005 | 765 (7.9%) | 747 (7.7%) | 0.630 | 0.007 |
| Neoplasms | 1,527 (15.2%) | 1,564 (15.5%) | 0.469 | 0.01 | 1,473 (15.3%) | 1,416 (14.7%) | 0.250 | 0.017 |
| **Mental health diagnoses** |  |  |  |  |  |  |  |  |
| Anxiety disorders | 950 (9.4%) | 974 (9.7%) | 0.565 | 0.008 | 924 (9.6%) | 900 (9.3%) | 0.482 | 0.010 |
| Major depressive disorders | 363 (3.6%) | 334 (3.3%) | 0.264 | 0.016 | 357 (3.7%) | 316 (3.3%) | 0.098 | 0.022 |
| PTSD and other trauma-related disorders | 156 (1.5%) | 164 (1.6%) | 0.652 | 0.006 | 148 (1.5%) | 137 (1.4%) | 0.512 | 0.009 |
| Bipolar disorder | 268 (2.7%) | 284 (2.8%) | 0.49 | 0.01 | 263 (2.7%) | 227 (2.4%) | 0.099 | 0.024 |
| Schizophrenia and other psychotic disorders | 231 (2.3%) | 243 (2.4%) | 0.577 | 0.008 | 222 (2.3%) | 186 (1.9%) | 0.072 | 0.026 |
| **Lifestyle-associated risk factors** |  |  |  |  |  |  |  |  |
| Nicotine dependence | 1,270 (12.6%) | 1,296 (12.9%) | 0.583 | 0.008 | 1,238 (12.8%) | 1,164 (12.1%) | 0.107 | 0.023 |
| Alcohol related disorders | 457 (4.5%) | 452 (4.5%) | 0.865 | 0.002 | 434 (4.5%) | 393 (4.1%) | 0.145 | 0.021 |
| Opioid related disorders | 206 (2.0%) | 184 (1.8%) | 0.261 | 0.016 | 198 (2.0%) | 193 (2.0%) | 0.798 | 0.004 |
| Cocaine related disorders | 314 (3.1%) | 308 (3.1%) | 0.807 | 0.003 | 303 (3.1%) | 262 (2.7%) | 0.080 | 0.025 |
| Cannabis related disorders | 326 (3.2%) | 310 (3.1%) | 0.519 | 0.009 | 318 (3.3%) | 278 (2.9%) | 0.096 | 0.024 |
| Polysubstance use disorders | 361 (3.6%) | 388 (3.9%) | 0.315 | 0.014 | 348 (3.6%) | 314 (3.3%) | 0.179 | 0.019 |

Data are presented as number (%) or mean ± standard deviation. P values reflect comparisons between matched cohorts and are provided for descriptive purposes only. Standardized differences $<0.10$ indicate negligible imbalance between groups.
Abbreviations: HBV, hepatitis B virus; HIV, human immunodeficiency virus; PTSD, post-traumatic stress disorder.

### Social Vulnerability Outcomes in HIV/HBV vs HIV Cohorts

In the matched HIV/HBV and HIV cohorts, median follow-up was 3.44 and 3.77 years, respectively (Table 2). Overall, the matched cohorts contributed 97,381 person-years of follow-up, including 45,622 in the HIV/HBV cohort and 51,759 in the HIV cohort. HIV/HBV coinfection was associated with a higher incidence of the primary social vulnerability composite compared with HIV monoinfection (11.5% vs 9.7%; IR, 2.50 vs 1.97 per 100 person-years; HR 1.25, 95% CI 1.15-1.37). At the domain level, HIV/HBV was associated with increased material vulnerability (HR 1.33, 95% CI 1.17-1.51) and healthcare access vulnerability (HR 1.29, 95% CI 1.16-1.44). Within material vulnerability, higher risks were observed for housing instability (HR 1.30, 95% CI 1.11-1.53), food insecurity (HR 1.32, 95% CI 1.04-1.69), and financial insecurity (HR 1.52, 95% CI 1.21-1.91). Within healthcare access, HIV/HBV was associated with insurance instability (HR 3.33, 95% CI 2.01-5.52) and care disengagement or nonadherence (HR 1.22, 95% CI 1.08-1.39). HIV/HBV was not associated with the interpersonal vulnerability or social environment composite, although social adjustment issues were modestly increased (HR 1.36, 95% CI 1.03-1.81).

**Table 2.** Incidence Rates and Hazard Ratios for Documented Social Vulnerability Outcomes in Adults With HIV/HBV Coinfection Compared With HIV Monoinfection.

| Outcomes | Cohorts |  |  | IR (95% CI)<br>(per 100 person-years) |  | HR (95% CI) | p-Value |
| --- | --- | --- | --- | --- | --- | --- | --- |
|  | Overall | HIV/HBV | HIV | HIV/HBV | HIV |  |  |
| **Follow up, Mean ± SD (years)** | 4.84 ± 4.43 | 4.53 ± 4.11 | 5.14 ± 4.76 | | | | |
| **Follow up, Median (IQR) (years)** | 3.60 (6.98) | 3.44 (6.28) | 3.77 (7.69) |  |  |  |  |
| **Primary outcome** |  |  |  |  |  |  |  |
| Social vulnerability composite | 18,278 | 1,028 (11.5%) | 909 (9.7%) | 2.50 (2.35–2.66) | 1.97 (1.85–2.10) | 1.25 (1.15–1.37) | **<0.001** |
| **Material vulnerability** |  |  |  |  |  |  |  |
| Material vulnerability composite | 19,436 | 529 (5.5%) | 437 (4.5%) | 1.18 (1.08–1.29) | 0.91 (0.83–1.00) | 1.33 (1.17–1.51) | **<0.001** |
| Housing instability | 19,615 | 344 (3.5%) | 287 (2.9%) | 0.76 (0.68–0.85) | 0.59 (0.53–0.67) | 1.30 (1.11–1.53) | **0.001** |
| Food insecurity | 20,058 | 144 (1.4%) | 121 (1.2%) | 0.31 (0.26–0.37) | 0.25 (0.21–0.30) | 1.32 (1.04–1.69) | **0.024** |
| Transportation insecurity | 20,093 | 80 (0.8%) | 65 (0.6%) | 0.17 (0.14–0.22) | 0.13 (0.10–0.17) | 1.35 (0.97–1.88) | 0.073 |
| Financial insecurity | 20,018 | 175 (1.7%) | 129 (1.3%) | 0.38 (0.32–0.44) | 0.26 (0.22–0.31) | 1.52 (1.21–1.91) | **<0.001** |
| Employment insecurity | 19,963 | 121 (1.2%) | 121 (1.2%) | 0.26 (0.22–0.31) | 0.25 (0.21–0.30) | 1.06 (0.82–1.37) | 0.650 |
| **Healthcare access and engagement** |  |  |  |  |  |  |  |
| Healthcare access composite | 18,924 | 721 (7.8%) | 603 (6.3%) | 1.66 (1.54–1.79) | 1.27 (1.17–1.38) | 1.29 (1.16–1.44) | **<0.001** |
| Insurance instability | 20,093 | 62 (0.6%) | 20 (0.2%) | 0.13 (0.10–0.17) | 0.04 (0.03–0.06) | 3.33 (2.01–5.52) | **<0.001** |
| Health access barriers | 20,091 | 59 (0.6%) | 55 (0.5%) | 0.13 (0.10–0.17) | 0.11 (0.09–0.15) | 1.18 (0.82–1.71) | 0.377 |
| Care disengagement or nonadherence | 16,863 | 508 (6.1%) | 468 (5.5%) | 1.32 (1.21–1.44) | 1.08 (0.99–1.19) | 1.22 (1.08–1.39) | **0.002** |
| **Interpersonal adversity** |  |  |  |  |  |  |  |
| Interpersonal adversity composite | 17,567 | 147 (1.7%) | 176 (2.0%) | 0.36 (0.30–0.42) | 0.39 (0.34–0.45) | 0.93 (0.74–1.16) | 0.501 |
| Adverse childhood experiences | 17,779 | 19 (0.2%) | 33 (0.4%) | 0.05 (0.03–0.07) | 0.07 (0.05–0.10) | 0.63 (0.36–1.11) | 0.104 |
| Primary support stressors | 17,701 | 101 (1.1%) | 129 (1.5%) | 0.25 (0.20–0.30) | 0.28 (0.24–0.34) | 0.88 (0.68–1.14) | 0.342 |
| Violence and victimization | 17,724 | 36 (0.4%) | 33 (0.4%) | 0.09 (0.06–0.12) | 0.07 (0.05–0.10) | 1.18 (0.73–1.89) | 0.499 |
| **Social vulnerability** |  |  |  |  |  |  |  |
| Social vulnerability composite | 17,540 | 222 (2.5%) | 226 (2.6%) | 0.55 (0.48–0.63) | 0.50 (0.44–0.57) | 1.16 (0.96–1.40) | 0.119 |
| Social isolation | 17,777 | 50 (0.6%) | 40 (0.4%) | 0.12 (0.09–0.16) | 0.09 (0.06–0.12) | 1.46 (0.96–2.22) | 0.076 |
| Social adjustment issues | 17,750 | 102 (1.2%) | 94 (1.1%) | 0.25 (0.20–0.30) | 0.21 (0.17–0.25) | 1.36 (1.03–1.81) | **0.032** |
| Legal and carceral issues | 17,760 | 44 (0.5%) | 55 (0.6%) | 0.11 (0.08–0.14) | 0.12 (0.09–0.15) | 0.95 (0.64–1.41) | 0.799 |

Incidence rates (IRs) and adjusted hazard ratios (HRs) are shown for incident social vulnerability outcomes among matched adults with HIV/HBV coinfection and HIV mono-infection. Analyses excluded individuals with the corresponding outcome before index.
Hazard ratios were estimated using Cox proportional hazards models adjusted for age, sex, race or ethnicity, calendar year of index, and baseline medical comorbidities. IRs are reported per 100 person-years. Follow-up is reported as mean ± SD and median (IQR).
The primary outcome was $\geq 1$ incident social vulnerability indicator after index.
Abbreviations: CI, confidence interval; HBV, hepatitis B virus; HIV, human immunodeficiency virus; HR, hazard ratio; IR, incidence rate; IQR, interquartile range; SD, standard deviation.

### Social Vulnerability Outcomes in HIV/HBV vs HBV Cohorts

In the matched HIV/HBV and HBV cohorts, median follow-up was 3.49 and 2.50 years, respectively (Table 3). Overall, the matched cohorts contributed 81,522 person-years of follow-up, including 44,335 in the HIV/HBV cohort and 37,187 in the HBV cohort. HIV/HBV coinfection was associated with substantially higher incidence of the primary social vulnerability composite compared with HBV monoinfection (11.0% vs 6.4%; IR, 2.39 vs 1.67 per 100 person-years; HR 1.50, 95% CI 1.35-1.67). The strongest and most consistent associations were observed for material vulnerability and healthcare access. HIV/HBV was associated with higher risk of the material vulnerability composite (HR 1.64, 95% CI 1.41-1.91), including housing instability (HR 1.94, 95% CI 1.59-2.36), food insecurity (HR 1.55, 95% CI 1.16-2.08), financial insecurity (HR 1.31, 95% CI 1.02-1.67), and employment insecurity (HR 1.46, 95% CI 1.07-1.98). HIV/HBV was also associated with the healthcare access composite (HR 1.61, 95% CI 1.41-1.83), driven primarily by care disengagement or nonadherence (HR 1.76, 95% CI 1.53-2.02). In contrast to the HIV comparison, HIV/HBV was associated with higher interpersonal vulnerability compared with HBV monoinfection (HR 1.45, 95% CI 1.13-1.85), driven by primary support stressors (HR 1.36, 95% CI 1.02-1.81) and violence or victimization (HR 2.16, 95% CI 1.23-3.79).

**Table 3.** Incidence Rates and Hazard Ratios for Documented Social Vulnerability Outcomes in Adults With HIV/HBV Coinfection Compared With HBV Monoinfection.

| Outcomes | Cohorts |  |  | IR (95% CI)<br>(per 100 person-years) |  | HR (95% CI) | p-Value |
| --- | --- | --- | --- | --- | --- | --- | --- |
|  | Overall | HIV/HBV | HBV | HIV/HBV | HBV |  |  |
| **Follow up, Mean ± SD (years)** | 4.22 ± 4.03 | 4.59 ± 4.14 | 3.85 ± 3.93 | | | | |
| **Follow up, Median (IQR) (years)** | 3.00 (6.01) | 3.49 (6.39) | 2.50 (5.63) |  |  |  |  |
| **Primary outcome** |  |  |  |  |  |  |  |
| Social vulnerability composite | 17,147 | 929 (11.0%) | 562 (6.4%) | 2.39 (2.24–2.55) | 1.67 (1.54–1.82) | 1.50 (1.35–1.67) | **<0.001** |
| **Material vulnerability** |  |  |  |  |  |  |  |
| Material vulnerability composite | 18,352 | 485 (5.3%) | 256 (2.8%) | 1.14 (1.04–1.25) | 0.71 (0.63–0.80) | 1.64 (1.41–1.91) | **<0.001** |
| Housing instability | 18,537 | 316 (3.4%) | 141 (1.5%) | 0.74 (0.66–0.83) | 0.39 (0.33–0.46) | 1.94 (1.59–2.36) | **<0.001** |
| Food insecurity | 18,917 | 128 (1.4%) | 69 (0.7%) | 0.30 (0.25–0.35) | 0.19 (0.15–0.24) | 1.55 (1.16–2.08) | **0.003** |
| Transportation insecurity | 18,955 | 68 (0.7%) | 52 (0.5%) | 0.16 (0.13–0.20) | 0.14 (0.11–0.19) | 1.11 (0.77–1.59) | 0.572 |
| Financial insecurity | 18,879 | 162 (1.7%) | 105 (1.1%) | 0.38 (0.32–0.44) | 0.29 (0.24–0.35) | 1.31 (1.02–1.67) | **0.033** |
| Employment insecurity | 18,831 | 111 (1.2%) | 65 (0.7%) | 0.26 (0.21–0.31) | 0.18 (0.14–0.23) | 1.46 (1.07–1.98) | **0.015** |
| **Healthcare access and engagement** |  |  |  |  |  |  |  |
| Healthcare access composite | 17,847 | 633 (7.2%) | 356 (3.9%) | 1.54 (1.42–1.67) | 0.99 (0.89–1.10) | 1.61 (1.41–1.83) | **<0.001** |
| Insurance instability | 18,933 | 58 (0.6%) | 40 (0.4%) | 0.13 (0.10–0.17) | 0.11 (0.08–0.15) | 1.21 (0.81–1.81) | 0.359 |
| Health access barriers | 18,941 | 51 (0.5%) | 37 (0.4%) | 0.12 (0.09–0.15) | 0.10 (0.07–0.14) | 1.18 (0.77–1.80) | 0.444 |
| Care disengagement or nonadherence | 18,243 | 598 (6.7%) | 311 (3.4%) | 1.47 (1.35–1.59) | 0.86 (0.77–0.96) | 1.76 (1.53–2.02) | **<0.001** |
| **Interpersonal adversity** |  |  |  |  |  |  |  |
| Interpersonal adversity composite | 19,029 | 172 (1.8%) | 103 (1.1%) | 0.40 (0.34–0.47) | 0.28 (0.23–0.34) | 1.45 (1.13–1.85) | **0.003** |
| Adverse childhood experiences | 19,259 | 22 (0.2%) | 20 (0.2%) | 0.05 (0.03–0.08) | 0.05 (0.03–0.08) | 0.96 (0.52–1.75) | 0.884 |
| Primary support stressors | 19,147 | 120 (1.3%) | 76 (0.8%) | 0.28 (0.23–0.33) | 0.21 (0.16–0.26) | 1.36 (1.02–1.81) | **0.038** |
| Violence and victimization | 19,232 | 42 (0.4%) | 17 (0.2%) | 0.10 (0.07–0.13) | 0.05 (0.03–0.07) | 2.16 (1.23–3.79) | **0.006** |
| **Social vulnerability** |  |  |  |  |  |  |  |
| Social vulnerability composite | 18,990 | 234 (2.5%) | 189 (2.0%) | 0.55 (0.48–0.63) | 0.52 (0.45–0.60) | 1.06 (0.88–1.29) | 0.526 |
| Social isolation | 19,265 | 57 (0.6%) | 41 (0.4%) | 0.13 (0.10–0.17) | 0.11 (0.08–0.15) | 1.18 (0.79–1.77) | 0.410 |
| Social adjustment issues | 19,224 | 105 (1.1%) | 75 (0.8%) | 0.24 (0.20–0.30) | 0.20 (0.16–0.25) | 1.19 (0.89–1.61) | 0.241 |
| Legal and carceral issues | 19,239 | 45 (0.5%) | 35 (0.4%) | 0.10 (0.08–0.14) | 0.09 (0.07–0.13) | 1.08 (0.69–1.68) | 0.733 |

Incidence rates (IRs) and adjusted hazard ratios (HRs) are shown for incident social vulnerability outcomes among matched adults with HIV/HBV coinfection and HBV monoinfection. Analyses excluded individuals with the corresponding outcome before index. Hazard ratios were estimated using Cox proportional hazards models adjusted for age, sex, race or ethnicity, calendar year of index, and baseline medical comorbidities. IRs are reported per 100 person-years. Follow-up is reported as mean ± SD and median (IQR). The primary outcome was $\geq 1$ incident social vulnerability indicator after index.
Abbreviations: CI, confidence interval; HBV, hepatitis B virus; HIV, human immunodeficiency virus; HR, hazard ratio; IR, incidence rate; IQR, interquartile range; SD, standard deviation.

### Sensitivity Analyses

Findings were robust across prespecified sensitivity analyses (Supplementary Tables S3-S8).

In the HIV comparison, 6-month lag analyses demonstrated persistent associations for the social vulnerability composite (HR 1.13, 95% CI 1.03-1.25), material vulnerability (HR 1.21, 95% CI 1.05-1.39), and healthcare access vulnerability (HR 1.21, 95% CI 1.07-1.37) (Supplementary Table S3). When separately adjusted for virologic markers and treatment, associations persisted for the social vulnerability composite (HR 1.29, 95% CI 1.15-1.44), material vulnerability (HR 1.50, 95% CI 1.29-1.75), and healthcare access vulnerability (HR 1.27, 95% CI 1.11-1.46) (Supplementary Table S4). When separately adjusted for healthcare utilization, associations remained significant for the social vulnerability composite (HR 1.21, 95% CI 1.12-1.31), material vulnerability (HR 1.15, 95% CI 1.01-1.30), and healthcare access vulnerability (HR 1.15, 95% CI 1.03-1.27) (Supplementary Table S5). Across all three analyses, insurance instability consistently showed the largest effect sizes (HRs 2.08-2.98).

For the HBV comparison, findings were highly consistent. Lag analyses demonstrated persistent associations for the social vulnerability composite (HR 1.50, 95% CI 1.33-1.68), material vulnerability (HR 1.80, 95% CI 1.51-2.13), healthcare access vulnerability (HR 1.55, 95% CI 1.34-1.79), and interpersonal vulnerability (HR 1.46, 95% CI 1.11-1.91) (Supplementary Table S6). In the model separately adjusted for virologic markers and treatment, associations persisted for the social vulnerability composite (HR 1.61, 95% CI 1.43-1.81), material vulnerability (HR 1.89, 95% CI 1.58-2.26), healthcare access vulnerability (HR 1.72, 95% CI 1.49-2.00), and interpersonal vulnerability (HR 1.41, 95% CI 1.08-1.84) (Supplementary Table S7). Finally, when separately adjusted for healthcare utilization, associations persisted for the social vulnerability composite (HR 1.75, 95% CI 1.58-1.95), material vulnerability (HR 1.81, 95% CI 1.56-2.10), healthcare access vulnerability (HR 1.84, 95% CI 1.62-2.09), and interpersonal vulnerability (HR 1.69, 95% CI 1.32-2.17) (Supplementary Table S8).

## DISCUSSION

In this large, propensity score-matched cohort study using federated EHR data from 110 U.S. healthcare organizations, HIV/HBV coinfection was independently associated with higher incidence of clinically documented social vulnerability compared with both HIV and HBV monoinfection. Excess risk was concentrated in material vulnerability and healthcare access disruption, with housing instability, food insecurity, financial insecurity, insurance instability, and care disengagement consistently elevated across both comparisons. The overall social vulnerability composite was elevated by 25% relative to HIV monoinfection and by 50% relative to HBV monoinfection, indicating that the social burden of coinfection exceeds what either infection alone would predict. Interpersonal adversity emerged only in the HBV comparison, revealing domain-specific patterns that vary by comparator. These associations persisted in 6-month landmark analyses, after adjustment for virologic markers and treatment variables, and after adjustment for healthcare utilization. These findings support the hypothesis that HIV/HBV coinfection is not merely a dual virologic state, but a social risk condition that amplifies the emergence of clinically recognized social adversity over time.

The consistent elevation of material vulnerability aligns with evidence that chronic multimorbidity produces cascading cycles of income loss, care rationing, and resource depletion [33]. Annual costs of first-line antiretroviral regimens in the United States range from $23,000 to $39,000, while Medicare Part D beneficiaries face $3,000 to $4,000 in annual out-of-pocket costs, which may contribute to dose-skipping and delayed refills that undermine viral suppression [33,34]. Adding HBV introduces additional monitoring, primary care and specialist visits, and the imperative for uninterrupted HBV-active therapy, which may further compound strain on individuals already at the margins of economic stability. The biological consequences may extend beyond treatment adherence. For instance, food insecurity has been independently associated with elevated immune activation markers (sCD14, sCD27, sCD163) and advanced liver fibrosis [35–37], and material deprivation has been associated with increased liver stiffness and a 3-fold higher risk of liver-related events in people with HIV [38]. In HIV/HBV, where both viruses contribute to hepatic inflammation, superimposed nutritional inadequacy may accelerate fibrosis independently of virologic control. That transportation and employment insecurity were not consistently elevated may reflect lower sensitivity of EHR documentation for these constructs; patient-reported measures may better capture the full spectrum of material hardship.

Healthcare access vulnerability was driven by care disengagement and, in the HIV comparison, a 3.3-fold higher risk of insurance instability. This carries particular clinical weight because any treatment gap in HIV/HBV risks HBV virologic rebound and hepatic flares, which occur in up to 51% of patients after nucleos(t)ide analogue cessation and can result in decompensation [39,40]. In the United States, health insurance coverage for people with HIV is fragmented across Medicaid, Medicare, Ryan White, and private insurance, with frequent transitions creating coverage gaps [33]. Financial barriers led to antiretroviral lapses in 10% of patients overall and 21% of those in AIDS Drug Assistance Programs (ADAPs) [41]. Unlike HIV, HBV monoinfection lacks any comparable safety-net infrastructure, and significant formulary barriers persist, with first-line antivirals frequently placed on high cost-sharing specialty tiers and one of the three first-line agents excluded by two of the largest pharmacy benefit managers in the United States [42]. The additional monitoring requirements of HBV coinfection may destabilize coverage beyond what HIV monoinfection alone produces. In this study, we found that care disengagement was consistently elevated across all analytic comparisons, consistent with evidence that structural life instability predicts psychosocial syndemic burden and worse virologic outcomes through pathways involving depression, reduced adherence, and detectable viral load [43]. Insurance coverage is directly linked to viral suppression: uninsured women without ADAP coverage had equivalent hazards of unsuppressed viral load compared with women insured by Medicaid [44]. These findings argue for integrated HIV/HBV care models with proactive insurance navigation to prevent coverage disruptions that cascade into treatment interruption. Beyond insurance navigation, integrated models should include structured SDoH screening and response pathways at predictable points of vulnerability, including diagnosis of coinfection, insurance transitions, ART changes, missed HBV-active therapy refills, missed hepatocellular carcinoma surveillance, and disengagement from HIV care.

We also found that interpersonal vulnerability was 45% higher in HIV/HBV coinfection relative to HBV monoinfection but did not differ from HIV monoinfection. People with HIV already carry disproportionate interpersonal adversity: approximately 87% report at least one adverse childhood experience, and more than one in four report lifetime intimate partner violence [45]. Adverse childhood experiences are associated with worse depressive symptoms, lower adherence, and reduced quality of life through pathways mediated by psychological distress [46]. The absence of a difference compared with HIV monoinfection likely reflects a ceiling effect. By contrast, people with HBV monoinfection in the United States are demographically distinct: approximately 70% were born outside the United States, with the highest prevalence among non-Hispanic Asian people, a group with different social risk profiles and potentially lower baseline documentation of interpersonal adversity [47,48]. Compared with HBV monoinfection, the 2.2-fold higher risk of violence or victimization and 1.4-fold higher primary support stressors in HIV/HBV consistent with the hypothesis that dual transmissible infections generate disclosure burden and relational strain. Among PWH, past-year intimate partner violence is associated with unstable housing, detectable viral load, and higher stigma scores [49]. These comparator-dependent patterns underscore the importance of domain-level analysis rather than reliance on composite measures alone.

Psychosocial vulnerability was not consistently elevated in HIV/HBV relative to either comparator. This may reflect the low sensitivity of EHR documentation for these constructs rather than genuine absence of risk. Social isolation and legal involvement are among the least reliably captured indicators, depending on patient disclosure and clinician inquiry [50]. Clinicians often lack training, time, and confidence in coding social risks, and documentation is frequently driven by billing incentives rather than systematic screening [50]. These domains may also be less visible during routine HIV or HBV care because they do not always present as immediate barriers to medication access, laboratory monitoring, or appointment completion. In the context of dual-infection stigma, where disclosure concerns may suppress both patient reporting and clinician inquiry, these constructs may be especially susceptible to documentation bias [50–52]. Thus, the null findings for this domain should be interpreted as evidence of limited EHR capture rather than proof that social isolation, legal stressors, or broader social-environmental instability are unimportant in HIV/HBV. Longitudinal cohorts with validated patient-reported outcome measures may better capture these dimensions.

A central contribution of this study is showing that SDoH can be analyzed not only as contextual exposures, but also as clinical signals that emerge during the course of HIV/HBV care. Area-level tools such as the CDC/ATSDR Social Vulnerability Index and Area Deprivation Index are useful for characterizing neighborhood disadvantage, but they do not reliably identify which individuals experience acute or recurrent social needs in clinical settings [25–28]. This distinction is important in HIV/HBV because the most relevant vulnerabilities may not be fixed attributes such as education or neighborhood deprivation, but time-sensitive disruptions in housing, food access, insurance coverage, and care continuity. The four-domain framework used here captures these clinically actionable disruptions as events that can appear, recur, cluster, and potentially resolve over time [29,30]. The association between HIV/HBV coinfection and newly documented SDoH, even after matching on 31 baseline variables, supports reframing social vulnerability as a measurable complication of complex chronic disease rather than a static background characteristic. Future studies should extend this framework by modeling SDoH longitudinally to determine whether dynamic social vulnerability better predicts downstream outcomes than baseline or area-level measures alone.

These findings may have implications for mechanistic inquiry and intervention design. Although this study examined SDoH documentation rather than downstream clinical outcomes, the concentration of excess risk in material vulnerability, healthcare access disruption, and interpersonal adversity suggests a plausible pathway through which newly emerging social vulnerability may later influence HIV/HBV morbidity. Chronic exposure to SDoH-driven psychosocial stress may activate the hypothalamic-pituitary-adrenal (HPA) axis, contributing to neuroendocrine dysregulation, immune activation, and allostatic load [53,54]. HIV itself can disrupt HPA regulation, potentially amplifying stress-related neuroendocrine and inflammatory pathways [55,56]. In HIV/HBV, material vulnerability and healthcare access disruption may therefore identify a subgroup at risk for downstream hepatic and extrahepatic complications through behavioral pathways, including treatment interruption and care disengagement, and through stress-biology pathways [57,58]. This hypothesis will require longitudinal studies linking dynamic SV to liver disease progression, extrahepatic comorbidity, and mechanistic biomarkers. The concentration of excess risk in material and healthcare access domains also identifies actionable interventions: housing support, food assistance, insurance navigation, and integrated care models address precisely the domains where coinfection confers greatest excess risk [59,60]. SDoH-focused hepatitis B interventions have demonstrated effectiveness in at least two-thirds of studies with comparative outcomes [51]. The dynamic framework can guide interventions responsive to episodic vulnerability rather than static risk classification.

This study has limitations. First, SDoH were identified using EHR documentation and ICD-10-CM Z codes, which have high specificity but limited sensitivity. Underascertainment is likely, particularly for stigma, social isolation, interpersonal violence, legal involvement, and other sensitive domains, and especially for HBV monoinfection, where these constructs are not routinely assessed in clinical management. Second, documentation may vary by health system, clinician screening practices, billing workflows, and patient willingness to disclose social risks. Therefore, the observed associations likely reflect clinically documented SDoH rather than the full burden of lived social vulnerability. Third, residual confounding remains possible despite propensity score matching and additional adjustment. Mental health, substance use, healthcare utilization, and social complexity may influence both exposure classification and SDoH documentation. Fourth, the EHR cannot fully distinguish new onset of social vulnerability from pre-existing vulnerability first documented after index. The 6-month lag analysis was designed to reduce this concern, but misclassification may persist. Fifth, HIV/HBV and comparator cohorts were defined using diagnosis codes and laboratory evidence, and misclassification of HBV chronicity, HIV status, or resolved infection is possible. Finally, this analysis focused on documented SDoH and did not directly evaluate subsequent hepatic, extrahepatic, or mortality outcomes; those relationships require separate longitudinal analyses. Despite these limitations, the strengths of this study include its large multi-institutional sample, dual-comparator design, rigorous propensity score matching, use of newly documented rather than prevalent SDoH outcomes, organization of outcomes within a prespecified dynamic individual-level framework, and robustness across lagged, virologic/treatment-adjusted, and healthcare utilization-adjusted sensitivity analyses.

In conclusion, HIV/HBV coinfection was associated with higher documentation of SDoH compared with both HIV and HBV monoinfection, with the most consistent excess risk in material vulnerability and healthcare access and engagement. These findings support a dynamic individual-level framework in which social vulnerability is not only a background characteristic but may emerge during the course of complex chronic viral coinfection. Recognizing SDoH as clinically trackable and potentially modifiable may help identify patients at higher risk for care disruption before social vulnerability translates into reduced monitoring continuity, treatment interruption, and downstream hepatic and extrahepatic morbidity.

## Data Availability

All data produced in the present work are contained in the manuscript

## AUTHOR CONTRIBUTIONS

GAY conceptualized the study with input from IO and KD. GAY curated the data and performed the statistical analyses. GAY, TC, AD, AA, AMM, CC, IO, and KD contributed to the development of the methods, analytic approach, and interpretation of findings. GAY secured funding and oversaw project administration. GAY drafted the original manuscript. All authors reviewed and revised the manuscript critically for important intellectual content and approved the final version. All authors had full access to the data. GAY and KD had final responsibility for the decision to submit the manuscript for publication.

## DATA SHARING

Deidentified individual participant data that underlie the results were extracted from TriNetX, a federated national health research network with data sourced from 110 health care organizations within the United States with waiver from WCG IRB.

## REFERENCES

1. Platt L, French CE, McGowan CR, Sabin K, Gower E, Trickey A, et al. Prevalence and burden of HBV co-infection among people living with HIV: a global systematic review and meta-analysis. J Viral Hepat. 2020;27(3):294–315. doi:10.1111/jvh.13217.

2. Bosh KA, Coyle JR, Hansen V, Kim EM, Speers S, Comer M, et al. HIV and viral hepatitis coinfection analysis using surveillance data from 15 US states and two cities. Epidemiol Infect. 2018;146(7):920–930. doi:10.1017/S0950268818000766.

3. Cheng Z, Lin P, Cheng N. HBV/HIV coinfection: impact on the development and clinical treatment of liver diseases. Front Med (Lausanne). 2021;8:713981. doi:10.3389/fmed.2021.713981.

4. Kim HN, Newcomb CW, Carbonari DM, Roy JA, Torgersen J, Althoff KN, et al. Risk of HCC with hepatitis B viremia among HIV/HBV-coinfected persons in North America. Hepatology. 2021;74(3):1190–1202. doi:10.1002/hep.31839.

5. Lo Re V 3rd, Newcomb CW, Carbonari DM, Roy JA, Althoff KN, Kitahata MM, et al. Determinants of liver complications among HIV/hepatitis B virus-coinfected patients. J Acquir Immune Defic Syndr. 2019;82(1):71–80. doi:10.1097/QAI.0000000000002094.

6. Menza TW, Hixson LK, Lipira L, Drach L. Social determinants of health and care outcomes among people with HIV in the United States. Open Forum Infect Dis. 2021;8(7):ofab330. doi:10.1093/ofid/ofab330.

7. Karram S, Sanger C, Convery C, Brantley A. Social determinants of health among persons living with HIV impact important health outcomes in Michigan. AIDS Behav. 2024;28(2):547–563. doi:10.1007/s10461-023-04243-5.

8. Schnarrs PW, Nash P, Ruderman SA, Crane HM, Delaney J, Singleton M, et al. The impact of childhood household violence on HIV-related outcomes in people with HIV in the United States: the role of psychological distress and social stressors. J Acquir Immune Defic Syndr. 2026;101(1):60–68. doi:10.1097/QAI.0000000000003772.

9. Hatcher AM, Smout EM, Turan JM, Christofides N, Stöckl H. Intimate partner violence and engagement in HIV care and treatment among women: a systematic review and meta-analysis. AIDS. 2015;29(16):2183–2194. doi:10.1097/QAD.0000000000000842.

10. Wong RJ, Telep LE, Wentworth CE, Liu Y, Iqbal S, Naik SD, et al. Hepatitis B virus treatment gaps in the US. JAMA Netw Open. 2025;8(11):e2542744. doi:10.1001/jamanetworkopen.2025.42744.

11. Ye Q, Kam LY, Yeo YH, Dang N, Huang DQ, Cheung R, et al. Substantial gaps in evaluation and treatment of patients with hepatitis B in the US. J Hepatol. 2022;76(1):63–74. doi:10.1016/j.jhep.2021.08.019.

12. Pham TTH, Toy M, Hutton D, Thompson W, Conners EE, Nelson NP, et al. Gaps and disparities in chronic hepatitis B monitoring and treatment in the United States, 2016-2019. Med Care. 2023;61(4):247–253. doi:10.1097/MLR.0000000000001825.

13. Ramier C, Boyd A, Smit C, van Zoest R, Claassen MAA, Pogány K, et al. Impact of socio-economic, behavioural and clinical factors on liver disease progression in individuals with HIV and hepatitis B. Liver Int. 2025;45(7):e70191. doi:10.1111/liv.70191.

14. Torres JM, Lawlor J, Colvin JD, Sills MR, Bettenhausen JL, Davidson A, et al. ICD social codes: an underutilized resource for tracking social needs. Med Care. 2017;55(9):810–816. doi:10.1097/MLR.0000000000000764.

15. Agarwal AR, Prichett L, Jain A, Srikumaran U. Assessment of use of ICD-9 and ICD-10 codes for social determinants of health in the US, 2011-2021. JAMA Netw Open. 2023;6(5):e2312538. doi:10.1001/jamanetworkopen.2023.12538.

16. Baker KM, Hill MA, Goldberg DG, Kitsantas P, Miller KE, Smith KM, et al. Using Z codes to document social risk factors in the electronic health record: a scoping review. Med Care. 2025;63(3):211–221. doi:10.1097/MLR.0000000000002101.

17. Aswani MS, Do LA, Shafer PR. Use of social determinants of health Z codes was sparse, 2016-22. Health Aff (Millwood). 2025;44(5):631–635. doi:10.1377/hlthaff.2024.01033.

18. Charkhchi P, Fazeli Dehkordy S, Carlos RC. Housing and food insecurity, care access, and health status among the chronically ill: an analysis of the Behavioral Risk Factor Surveillance System. J Gen Intern Med. 2018;33(5):644–650. doi:10.1007/s11606-017-4255-z.

19. Sloan CE, Lourie MA, Bowling CB, Pignone M, Van Houtven CH, McDermott CL. A patient-informed framework of financial strain among adults with multimorbidity. J Gen Intern Med. 2026;41(3):744–752. doi:10.1007/s11606-025-09911-x.

20. Russ LW, Meyer AC, Takahashi LM, Ou S, Tran J, Cruz P, et al. Examining barriers to care: provider and client perspectives on the stigmatization of HIV-positive Asian Americans with and without viral hepatitis co-infection. AIDS Care. 2012;24(10):1302–1307. doi:10.1080/09540121.2012.658756.

21. Yendewa GA, Sellu EJ, Kpaka RA, James PB, Yendewa SA, Cummings PE, et al. Measuring stigma associated with hepatitis B virus infection in Sierra Leone: validation of an abridged Berger HIV stigma scale. J Viral Hepat. 2023;30(7):621–629. doi:10.1111/jvh.13838.

22. Lekas HM, Siegel K, Leider J. Felt and enacted stigma among HIV/HCV-coinfected adults: the impact of stigma layering. Qual Health Res. 2011;21(9):1205–1219. doi:10.1177/1049732311405684.

23. Singer M, Bulled N, Ostrach B, Mendenhall E. Syndemics and the biosocial conception of health. Lancet. 2017;389(10072):941–950. doi:10.1016/S0140-6736(17)30003-X.

24. Xiao Z, Li Z, Chen Q. Socio-economic and geographic factors in liver disease progression among individuals with HIV-HBV coinfection. Liver Int. 2025;45(9):e70249. doi:10.1111/liv.70249.

25. Commission on Social Determinants of Health. Closing the Gap in a Generation: Health Equity Through Action on the Social Determinants of Health. Geneva: World Health Organization; 2008.

26. Office of Disease Prevention and Health Promotion. Healthy People 2030: Social determinants of health. Washington (DC): U.S. Department of Health and Human Services; 2020.

27. Kind AJ, Jencks S, Brock J, Yu M, Bartels C, Ehlenbach W, et al. Neighborhood socioeconomic disadvantage and 30-day rehospitalization: a retrospective cohort study. Ann Intern Med. 2014;161(11):765–774. doi:10.7326/M13-2946.

28. Brown EM, Franklin SM, Ryan JL, Canterberry M, Bowe A, Pantell MS, et al. Assessing area-level deprivation as a proxy for individual-level social risks. Am J Prev Med. 2023;65(6):1163–1171. doi:10.1016/j.amepre.2023.06.006.

29. Rajabiun S, Davis-Plourde K, Tinsley M, Quinn EK, Borne D, Maskay MH, et al. Pathways to housing stability and viral suppression for people living with HIV/AIDS: findings from the Building a Medical Home for Multiply Diagnosed HIV-positive Homeless Populations initiative. PLoS One. 2020;15(10):e0239190. doi:10.1371/journal.pone.0239190.

30. Bleasdale J, Liu Y, Leone LA, Morse GD, Przybyla SM. The impact of food insecurity on receipt of care, retention in care, and viral suppression among people living with HIV/AIDS in the United States: a causal mediation analysis. Front Public Health. 2023;11:1133328. doi:10.3389/fpubh.2023.1133328.

31. Singal AG, Llovet JM, Yarchoan M, Mehta N, Heimbach JK, Dawson LA, et al. AASLD Practice Guidance on prevention, diagnosis, and treatment of hepatocellular carcinoma. Hepatology. 2023;78(6):1922–1965. doi:10.1097/HEP.0000000000000466.

32. Rich JD, Beckwith CG, Macmadu A, Marshall BDL, Brinkley-Rubinstein L, Amon JJ, et al. Clinical care of incarcerated people with HIV, viral hepatitis, or tuberculosis. Lancet. 2016;388(10049):1103–1114. doi:10.1016/S0140-6736(16)30379-8.

33. Kates J, Dawson L, Horn TH, Killelea A, McCann NC, Crowley JS, et al. Insurance coverage and financing landscape for HIV treatment and prevention in the USA. Lancet. 2021;397(10279):1127–1138. doi:10.1016/S0140-6736(21)00397-4.

34. Tseng CW, Dudley RA, Chen R, Walensky RP. Medicare Part D and cost-sharing for antiretroviral therapy and preexposure prophylaxis. JAMA Netw Open. 2020;3(4):e202739. doi:10.1001/jamanetworkopen.2020.2739.

35. Valerio LA, Rzepka MC, Davy-Mendez T, Williams A, Perhac A, Napravnik S, et al. Food insecurity prevalence and risk factors among persons with HIV in a Southeastern US clinical care setting. AIDS Behav. 2025;29(1):45–54. doi:10.1007/s10461-024-04497-7.

36. Tamargo JA, Hernandez-Boyer J, Teeman C, Martin HR, Huang Y, Johnson A, et al. Immune activation: a link between food insecurity and chronic disease in people living with human immunodeficiency virus. J Infect Dis. 2021;224(12):2043–2052. doi:10.1093/infdis/jiab257.

37. Tamargo JA, Sherman KE, Campa A, Martinez SS, Li T, Hernandez J, et al. Food insecurity is associated with magnetic resonance-determined nonalcoholic fatty liver and liver fibrosis in low-income, middle-aged adults with and without HIV. Am J Clin Nutr. 2021;113(3):593–601. doi:10.1093/ajcn/nqaa362.

38. Long C, Cinque F, Kablawi D, Kardashian A, Lloyd A, Vilar-Gomez E, et al. Material deprivation is associated with liver stiffness and liver-related outcomes in people with HIV. Liver Int. 2024.

39. Dongelmans EJ, Hirode G, Hansen BE, Chen CH, Su TH, Seto WK, et al. Predictors of hepatic flares after nucleos(t)ide analogue cessation – Results of a global cohort study (RETRACT-B study). J Hepatol. 2025;82(3):446–455. doi:10.1016/j.jhep.2024.08.015.

40. Chang ML, Liaw YF. Hepatitis B flares in chronic hepatitis B: pathogenesis, natural course, and management. J Hepatol. 2014;61(6):1407–1417. doi:10.1016/j.jhep.2014.08.033.

41. Wohl DA, Kuwahara RK, Javadi K, Kirby C, Rosen DL, Napravnik S, et al. Financial barriers and lapses in treatment and care of HIV-infected adults in a Southern state in the United States. AIDS Patient Care STDS. 2017;31(11):463–469. doi:10.1089/apc.2017.0125.

42. Hepatitis B Foundation. Drug tiering of hepatitis B antivirals: a consumer report. Plymouth Meeting (PA): Hepatitis B Foundation; 2020. Available from: https://www.hepb.org/assets/Uploads/Drug-Tiering-Consumer-Report-10.27.2020-Final-1-compressed.pdf (accessed on May 30, 2026).

43. Weinstein ER, Kirakosian N, Chen YO, Bharat B, Safren SA. Life instability and its effects on psychosocial syndemic problems and HIV-care outcomes in people living with HIV in care in South Florida. J Acquir Immune Defic Syndr. 2026;101(2):120–127. doi:10.1097/QAI.0000000000003777.

44. Ludema C, Cole SR, Eron JJ Jr, Edmonds A, Holmes GM, Anastos K, et al. Impact of health insurance, ADAP, and income on HIV viral suppression among US women in the Women’s Interagency HIV Study, 2006-2009. J Acquir Immune Defic Syndr. 2016;73(3):307–312. doi:10.1097/QAI.0000000000001078.

45. Sanders R, Dombrowski JC, Hajat A, Buskin S, Erly S. Associations between adverse childhood experiences, viral suppression, and quality of life among persons living with HIV in Washington state. AIDS Care. 2024;36(7):937–945. doi:10.1080/09540121.2023.2299339.

46. Young-Wolff KC, Sarovar V, Sterling SA, Leibowitz A, McCaw B, Hare CB, et al. Adverse childhood experiences, mental health, substance use, and HIV-related outcomes among persons with HIV. AIDS Care. 2019;31(10):1241–1249. doi:10.1080/09540121.2019.1587372.

47. Roberts H, Ly KN, Yin S, Hughes E, Teshale E, Jiles R. Prevalence of HBV infection, vaccine-induced immunity, and susceptibility among at-risk populations: US households, 2013-2018. Hepatology. 2021;74(5):2353–2365. doi:10.1002/hep.31991.

48. Le MH, Yeo YH, Cheung R, Henry L, Lok AS, Nguyen MH. Chronic hepatitis B prevalence among foreign-born and U.S.-born adults in the United States, 1999-2016. Hepatology. 2020;71(2):431–443. doi:10.1002/hep.30831.

49. Fredericksen RJ, Mixson LS, Drumright LN, Nance RM, Delaney JAC, Ruderman SA, et al. Correlates of intimate partner violence, including psychological partner violence, in a multisite U.S. cohort of people in HIV care. AIDS Behav. 2024;28(9):3170–3183. doi:10.1007/s10461-024-04402-2.

50. Enich M, Tiderington E. Physician perspectives on Z codes for social determinants of health screening. J Gen Intern Med. 2026;41(2):391–398. doi:10.1007/s11606-025-09548-w.

51. Anyiwe K, Erman A, Hassan M, Feld JJ, Pullenayegum E, Wong WWL, et al. Characterising the effectiveness of social determinants of health-focused hepatitis B interventions: a systematic review. Lancet Infect Dis. 2024;24(6):e366–e385. doi:10.1016/S1473-3099(23)00590-X.

52. Hyun S, Ko O, Kim S, Ventura WR. Sociocultural barriers to hepatitis B health literacy in an immigrant population: a focus group study in Korean Americans. BMC Public Health. 2021;21(1):404. doi:10.1186/s12889-021-10441-4.

53. Russell G, Lightman S. The human stress response. Nat Rev Endocrinol. 2019;15(9):525–534. doi:10.1038/s41574-019-0228-0.

54. Selvarajah S, Corona Maioli S, Deivanayagam TA, de Morais Sato P, Devakumar D, Kim SS, et al. Racism, xenophobia, and discrimination: mapping pathways to health outcomes. Lancet. 2022;400(10368):2109–2124. doi:10.1016/S0140-6736(22)02484-9.

55. George MM, Bhangoo A. Human immune deficiency virus (HIV) infection and the hypothalamic pituitary adrenal axis. Rev Endocr Metab Disord. 2013;14(2):105–112. doi:10.1007/s11154-013-9244-x.

56. Chrousos GP, Zapanti ED. Hypothalamic-pituitary-adrenal axis in HIV infection and disease. Endocrinol Metab Clin North Am. 2014;43(3):791–806. doi:10.1016/j.ecl.2014.06.002.

57. Crane M, Avihingsanon A, Rajasuriar R, Velayudham P, Iser D, Solomon A, et al. Lipopolysaccharide, immune activation, and liver abnormalities in HIV/hepatitis B virus (HBV)-coinfected individuals receiving HBV-active combination antiretroviral therapy. J Infect Dis. 2014;210(5):745–751. doi:10.1093/infdis/jiu119.

58. Xu M, Warner C, Duan X, Cheng Z, Jeyarajan AJ, Li W, et al. HIV coinfection exacerbates HBV-induced liver fibrogenesis through a HIF-1α- and TGF-β1-dependent pathway. J Hepatol. 2024;80(6):868–881. doi:10.1016/j.jhep.2024.01.026.

59. Aidala AA, Wilson MG, Shubert V, Gogolishvili D, Globerman J, Rueda S, et al. Housing status, medical care, and health outcomes among people living with HIV/AIDS: a systematic review. Am J Public Health. 2016;106(1):e1–e23. doi:10.2105/AJPH.2015.302905.

60. Dombrowski JC, Corcorran MA, Carney T, Parczewski M, Gandhi M. The impact of homelessness and housing insecurity on HIV. Lancet HIV. 2025;12(6):e449–e458. doi:10.1016/S2352-3018(25)00048-7.

