## Supplementary Tables for "Social Determinants of Health in HIV/HBV Coinfection Compared with HIV and HBV Monoinfection: A Framework for Dynamic Individual-Level Social Vulnerability"

**Supplementary Table S1.** Baseline Characteristics Before and After Propensity Score Matching for Adults With HIV/HBV Coinfection Compared With HIV Monoinfection

| Variables | Before Matching | | | | | After Matching | | | |
| --- | --- | --- | --- | --- | --- | --- | --- | --- | --- |
|  | **HIV/HBV** | **HIV** | **p-Value** | **Standard difference** | **HIV/HBV** | | **HIV** | **p-Value** | **Standard difference** |
| Totals | 10,080 | 279,283 |  |  | 10,071 | | 10,071 |  |  |
| Age at Index (years) | 47.1 ± 12.9 | 43.5 ± 14.4 | **<0.001** | 0.261 | 47.1 ± 12.9 | | 47.3 ± 13.8 | 0.246 | 0.016 |
| Gender |  |  |  |  |  | |  |  |  |
| Male | 7,993 (79.3%) | 203,869 (73.0%) | **<0.001** | 0.148 | 7,984 (79.3%) | | 7,935 (78.8%) | 0.080 | 0.012 |
| Female | 2,082 (20.7%) | 75,177 (26.9%) | **<0.001** | 0.147 | 2,082 (20.7%) | | 2,151 (21.4%) | 0.230 | 0.018 |
| Race or ethnicity |  |  |  |  |  | |  |  |  |
| White | 3,371 (33.4%) | 105,623 (37.8%) | **<0.001** | 0.091 | 3,368 (33.4%) | | 3,488 (34.6%) | 0.074 | 0.025 |
| Asian | 417 (4.1%) | 6,893 (2.5%) | **<0.001** | 0.093 | 417 (4.1%) | | 421 (4.2%) | 0.888 | 0.002 |
| Unknown Race | 1,597 (15.8%) | 36,410 (13.0%) | **<0.001** | 0.080 | 1,596 (15.8%) | | 1,628 (16.2%) | 0.539 | 0.009 |
| Black or African American | 4,342 (43.1%) | 112,564 (40.3%) | **<0.001** | 0.056 | 4,337 (43.1%) | | 4,230 (42.0%) | 0.120 | 0.022 |
| Hispanic or Latino | 477 (4.7%) | 27,578 (9.9%) | **<0.001** | 0.199 | 477 (4.7%) | | 473 (4.7%) | 0.894 | 0.002 |
| Comorbidities |  |  |  |  |  | |  |  |  |
| Hypertensive diseases | 2,246 (22.3%) | 22,000 (7.9%) | **<0.001** | 0.411 | 2,237 (22.2%) | | 2,320 (23.0%) | 0.091 | 0.038 |
| Ischemic heart diseases | 566 (5.6%) | 5,651 (2.0%) | **<0.001** | 0.188 | 562 (5.6%) | | 564 (5.6%) | 0.951 | 0.001 |
| Heart failure | 310 (3.1%) | 2,705 (1.0%) | **<0.001** | 0.150 | 308 (3.1%) | | 292 (2.9%) | 0.507 | 0.009 |
| Diabetes mellitus | 869 (8.6%) | 9,903 (3.5%) | **<0.001** | 0.214 | 864 (8.6%) | | 919 (9.1%) | 0.172 | 0.019 |
| Chronic lower respiratory diseases | 1,060 (10.5%) | 13,729 (4.9%) | **<0.001** | 0.211 | 1,054 (10.5%) | | 1,101 (10.9%) | 0.284 | 0.015 |
| Chronic kidney disease | 743 (7.4%) | 4,605 (1.6%) | **<0.001** | 0.278 | 736 (7.3%) | | 705 (7.0%) | 0.397 | 0.012 |
| Diseases of liver | 797 (7.9%) | 4,112 (1.5%) | **<0.001** | 0.308 | 790 (7.8%) | | 803 (8.0%) | 0.734 | 0.005 |
| Neoplasms | 1,533 (15.2%) | 15,511 (5.6%) | **<0.001** | 0.321 | 1,527 (15.2%) | | 1,564 (15.5%) | 0.469 | 0.01 |
| Mental health diagnoses |  |  |  |  |  | |  |  |  |
| Anxiety disorders | 956 (9.5%) | 11,734 (4.2%) | **<0.001** | 0.210 | 950 (9.4%) | | 974 (9.7%) | 0.565 | 0.008 |
| Major depressive disorders | 366 (3.6%) | 3,817 (1.4%) | **<0.001** | 0.145 | 363 (3.6%) | | 334 (3.3%) | 0.264 | 0.016 |
| PTSD and trauma-related disorders | 157 (1.6%) | 2,389 (0.9%) | **<0.001** | 0.064 | 156 (1.5%) | | 164 (1.6%) | 0.652 | 0.006 |
| Bipolar disorder | 269 (2.7%) | 3,914 (1.4%) | **<0.001** | 0.090 | 268 (2.7%) | | 284 (2.8%) | 0.490 | 0.01 |
| Schizophrenia and other psychotic disorders | 233 (2.3%) | 3,311 (1.2%) | **<0.001** | 0.086 | 231 (2.3%) | | 243 (2.4%) | 0.577 | 0.008 |
| Lifestyle-associated risk factors |  |  |  |  |  | |  |  |  |
| Nicotine dependence | 1,277 (12.7%) | 14,042 (5.0%) | **<0.001** | 0.272 | 1,270 (12.6%) | | 1,296 (12.9%) | 0.583 | 0.008 |
| Alcohol related disorders | 459 (4.6%) | 5,571 (2.0%) | **<0.001** | 0.144 | 457 (4.5%) | | 452 (4.5%) | 0.865 | 0.002 |
| Opioid related disorders | 207 (2.1%) | 2,181 (0.8%) | **<0.001** | 0.108 | 206 (2.0%) | | 184 (1.8%) | 0.261 | 0.016 |
| Cocaine related disorders | 318 (3.2%) | 2,913 (1.0%) | **<0.001** | 0.148 | 314 (3.1%) | | 308 (3.1%) | 0.807 | 0.003 |
| Cannabis related disorders | 330 (3.3%) | 3,453 (1.2%) | **<0.001** | 0.138 | 326 (3.2%) | | 310 (3.1%) | 0.519 | 0.009 |
| Polysubstance use disorders | 364 (3.6%) | 3,783 (1.4%) | **<0.001** | 0.145 | 361 (3.6%) | | 388 (3.9%) | 0.315 | 0.014 |
| Healthcare utilization |  |  |  |  |  | |  |  |  |
| Ambulatory | 8,692 (85.2%) | 209,036 (73.5%) | **<0.001** | 0.294 | 7,251 (84.2%) | | 7,277 (84.5%) | 0.585 | 0.008 |
| Emergency department | 4,082 (40.0%) | 91,585 (32.2%) | **<0.001** | 0.164 | 3,258 (37.8%) | | 3,168 (36.8%) | 0.156 | 0.022 |
| Inpatient | 4,379 (42.9%) | 67,278 (23.6%) | **<0.001** | 0.418 | 3,424 (39.8%) | | 3,494 (40.6%) | 0.277 | 0.017 |
| Medications |  |  |  |  |  | |  |  |  |
| NRTIs | 4,676 (55.4%) | 57,418 (26.3%) | **<0.001** | 0.618 | 4,387 (53.8%) | | 4,267 (52.3%) | 0.060 | 0.029 |
| Emtricitabine | 3,797 (45.0%) | 43,764 (20.1%) | **<0.001** | 0.551 | 3,594 (44.1%) | | 3,538 (43.4%) | 0.377 | 0.014 |
| Tenofovir disoproxil | 3,110 (36.8%) | 29,064 (13.3%) | **<0.001** | 0.563 | 2,911 (35.7%) | | 2,853 (35.0%) | 0.342 | 0.015 |
| Tenofovir alafenamide | 2,496 (29.6%) | 21,672 (9.9%) | **<0.001** | 0.508 | 2,328 (28.5%) | | 2,281 (28.0%) | 0.414 | 0.013 |
| Lamivudine | 1,314 (15.6%) | 14,067 (6.5%) | **<0.001** | 0.294 | 1,162 (14.2%) | | 1,105 (13.5%) | 0.197 | 0.02 |
| Entecavir | 262 (2.6%) | - | - | - | 223 (2.6%) | | - | - | - |
| NNRTIs | 1,474 (17.5%) | 17,261 (7.9%) | **<0.001** | 0.29 | 1,363 (16.7%) | | 1,328 (16.3%) | 0.460 | 0.012 |
| PIs | 1,754 (20.8%) | 17,099 (7.8%) | **<0.001** | 0.376 | 1,600 (19.6%) | | 1,588 (19.5%) | 0.813 | 0.004 |
| INSTIs | 2,222 (26.3%) | 21,025 (9.6%) | **<0.001** | 0.445 | 2,031 (24.9%) | | 1,981 (24.3%) | 0.363 | 0.014 |
| HIV combinations | 1,832 (21.7%) | 16,089 (7.4%) | **<0.001** | 0.415 | 1,696 (20.8%) | | 1,667 (20.4%) | 0.575 | 0.009 |
| Laboratory markers |  |  |  |  |  | |  |  |  |
| CD4 count (cells/μL) |  |  |  |  |  | |  |  |  |
| Median (IQR) | 396 (483) | 476 (497) | **<0.001** | 0.156 | 398 (484) | | 425 (548) | 0.063 | 0.162 |
| Available, n (%) | 3,841 (41.9%) | 37,388 (15.5%) |  |  | 3,775 (41.9%) | | 3,655 (40.5%) |  |  |
| CD4 percentage (%) |  |  |  |  |  | |  |  |  |
| Median (IQR) | 24 (20) | 28 (19) | **<0.001** | 0.124 | 24 (20) | | 27 (20) | 0.654 | 0.078 |
| Available, n (%) | 4,272 (46.6%) | 41,220 (17.1%) |  |  | 4,124 (45.8%) | | 4,094 (45.4%) |  |  |
| HIV RNA (copies/mL) |  |  |  |  |  | |  |  |  |
| Median (IQR) | 2,080 (52,111) | 1,500 (40,510) | **<0.001** | 0.080 | 2,324 (53,235) | | 704 (31,441) | 0.066 | 0.097 |
| Available, n (%) | 2,049 (22.4%) | 15,932 (6.6%) |  |  | 1,994 (22.1%) | | 1,908 (21.2%) |  |  |
| HBV DNA (IU/mL) |  |  |  |  |  | |  |  |  |
| Median (IQR) | 190 (3,880) |  |  |  | 181 (3,020) | |  |  |  |
| Available, n (%) | 524 (5.7%) |  |  |  | 508 (5.6%) | |  |  |  |

Baseline demographic, clinical, mental health, lifestyle‑associated characteristics, healthcare utilization, antiretroviral medications, and laboratory markers are shown before and after propensity score matching for adults with HIV/HBV coinfection versus HIV monoinfection. Propensity score matching was performed using 1:1 nearest‑neighbor matching without replacement. Data are presented as number (%) or mean ± standard deviation. P‑values are provided for descriptive purposes only. Standardized differences <0.10 indicate negligible imbalance between groups.

Laboratory availability is shown for the baseline period (≤6 months before index). Extending the lookback window would increase capture; thus, these percentages are minimum estimates and should not be interpreted as the true testing frequency in the cohorts.

Abbreviations: HBV, hepatitis B virus; HIV, human immunodeficiency virus; NRTI, nucleoside/nucleotide reverse transcriptase inhibitor; NNRTI, non‑nucleoside reverse transcriptase inhibitor; PI, protease inhibitor; INSTI, integrase strand transfer inhibitor; IQR, interquartile range.

**Supplementary Table S2.** Baseline Characteristics Before and After Propensity Score Matching for Adults With HIV/HBV Coinfection Compared With HBV Monoinfection

| Variables | Before Matching | | | | | After Matching | | | |
| --- | --- | --- | --- | --- | --- | --- | --- | --- | --- |
|  | **HIV/HBV** | **HBV** | **p-Value** | **Standard difference** | **HIV/HBV** | | **HBV** | **p-Value** | **Standard difference** |
| Totals | 9,663 | 426,955 |  |  | 9,659 | | 9,659 |  |  |
| Age at Index (years) | 47.0 ± 12.9 | 46.9 ± 17.6 | 0.559 | 0.007 | 47.0 ± 12.9 | | 47.4 ± 14.3 | 0.074 | 0.026 |
| Gender |  |  |  |  |  | |  |  |  |
| Male | 7,643 (79.1%) | 177,903 (41.7%) | **<0.001** | 0.828 | 7,639 (79.1%) | | 7,680 (79.5%) | 0.080 | 0.010 |
| Female | 2,015 (20.9%) | 248,889 (58.3%) | **<0.001** | 0.829 | 2,015 (20.9%) | | 1,979 (20.5%) | 0.367 | 0.018 |
| Race or ethnicity |  |  |  |  |  | |  |  |  |
| White | 3,281 (34.0%) | 117,064 (27.4%) | **<0.001** | 0.142 | 3,281 (34.0%) | | 3,345 (34.6%) | 0.332 | 0.014 |
| Asian | 410 (4.2%) | 99,938 (23.4%) | **<0.001** | 0.578 | 410 (4.2%) | | 408 (4.2%) | 0.943 | 0.001 |
| Unknown Race | 1,574 (16.3%) | 157,705 (36.9%) | **<0.001** | 0.481 | 1,574 (16.3%) | | 1,563 (16.2%) | 0.830 | 0.003 |
| Black or African American | 4,073 (42.2%) | 41,259 (9.7%) | **<0.001** | 0.798 | 4,069 (42.1%) | | 3,998 (41.4%) | 0.300 | 0.015 |
| Hispanic or Latino | 467 (4.8%) | 5,581 (1.3%) | **<0.001** | 0.205 | 467 (4.8%) | | 500 (5.2%) | 0.276 | 0.016 |
| Comorbidities |  |  |  |  |  | |  |  |  |
| Hypertensive diseases | 2,142 (22.2%) | 58,989 (13.8%) | **<0.001** | 0.219 | 2,140 (22.2%) | | 2,181 (22.6%) | 0.479 | 0.010 |
| Ischemic heart diseases | 540 (5.6%) | 18,753 (4.4%) | **<0.001** | 0.055 | 540 (5.6%) | | 538 (5.6%) | 0.950 | 0.001 |
| Heart failure | 293 (3.0%) | 9,304 (2.2%) | **<0.001** | 0.054 | 293 (3.0%) | | 299 (3.1%) | 0.802 | 0.004 |
| Diabetes mellitus | 844 (8.7%) | 32,393 (7.6%) | **<0.001** | 0.042 | 844 (8.7%) | | 867 (9.0%) | 0.560 | 0.008 |
| Chronic lower respiratory diseases | 1,008 (10.4%) | 26,104 (6.1%) | **<0.001** | 0.157 | 1,007 (10.4%) | | 935 (9.7%) | 0.085 | 0.025 |
| Chronic kidney disease | 712 (7.4%) | 18,349 (4.3%) | **<0.001** | 0.131 | 711 (7.4%) | | 687 (7.1%) | 0.505 | 0.010 |
| Diseases of liver | 765 (7.9%) | 25,000 (5.9%) | **<0.001** | 0.081 | 765 (7.9%) | | 747 (7.7%) | 0.630 | 0.007 |
| Neoplasms | 1,476 (15.3%) | 55,365 (13.0%) | **<0.001** | 0.066 | 1,473 (15.3%) | | 1,416 (14.7%) | 0.250 | 0.017 |
| Mental health diagnoses |  |  |  |  |  | |  |  |  |
| Anxiety disorders | 927 (9.6%) | 23,408 (5.5%) | **<0.001** | 0.156 | 924 (9.6%) | | 900 (9.3%) | 0.482 | 0.010 |
| Major depressive disorders | 358 (3.7%) | 4,435 (1.0%) | **<0.001** | 0.176 | 357 (3.7%) | | 316 (3.3%) | 0.098 | 0.022 |
| PTSD and other trauma-related disorders | 149 (1.5%) | 1,690 (0.4%) | **<0.001** | 0.117 | 148 (1.5%) | | 137 (1.4%) | 0.512 | 0.009 |
| Bipolar disorder | 265 (2.7%) | 3,357 (0.8%) | **<0.001** | 0.149 | 263 (2.7%) | | 227 (2.4%) | 0.099 | 0.024 |
| Schizophrenia and other psychotic disorders | 223 (2.3%) | 3,077 (0.7%) | **<0.001** | 0.13 | 222 (2.3%) | | 186 (1.9%) | 0.072 | 0.026 |
| Lifestyle-associated risk factors |  |  |  |  |  | |  |  |  |
| Nicotine dependence | 1,242 (12.9%) | 13,825 (3.2%) | **<0.001** | 0.359 | 1,238 (12.8%) | | 1,164 (12.1%) | 0.107 | 0.023 |
| Alcohol related disorders | 435 (4.5%) | 5,421 (1.3%) | **<0.001** | 0.194 | 434 (4.5%) | | 393 (4.1%) | 0.145 | 0.021 |
| Opioid related disorders | 198 (2.0%) | 1,882 (0.4%) | **<0.001** | 0.145 | 198 (2.0%) | | 193 (2.0%) | 0.798 | 0.004 |
| Cocaine related disorders | 307 (3.2%) | 1,361 (0.3%) | **<0.001** | 0.219 | 303 (3.1%) | | 262 (2.7%) | 0.080 | 0.025 |
| Cannabis related disorders | 320 (3.3%) | 2,399 (0.6%) | **<0.001** | 0.201 | 318 (3.3%) | | 278 (2.9%) | 0.096 | 0.024 |
| Polysubstance use disorders | 350 (3.6%) | 2,673 (0.6%) | **<0.001** | 0.209 | 348 (3.6%) | | 314 (3.3%) | 0.179 | 0.019 |
| Healthcare utilization |  |  |  |  |  | |  |  |  |
| Ambulatory | 8,542 (85.1%) | 375,642 (87.0%) | **<0.001** | 0.056 | 6,617 (83.6%) | | 6,596 (83.3%) | 0.653 | 0.007 |
| Emergency department | 4,003 (39.9%) | 132,243 (30.6%) | **<0.001** | 0.194 | 2,980 (37.7%) | | 3,006 (38.0%) | 0.670 | 0.007 |
| Inpatient | 4,280 (42.6%) | 162,759 (37.7%) | **<0.001** | 0.101 | 3,211 (40.6%) | | 3,188 (40.3%) | 0.710 | 0.006 |
| Medications |  |  |  |  |  | |  |  |  |
| **NRTIs** | 5,417 (54.0%) | 30,202 (7.0%) | **<0.001** | 1.187 | 3,295 (41.6%) | | 2,916 (36.8%) | **<0.001** | 0.098 |
| Emtricitabine | 4,412 (44.0%) | - | - | - | 2,616 (33.1%) | | - | - | - |
| Tenofovir disoproxil | 3,601 (35.9%) | 7,043 (1.6%) | **<0.001** | 0.976 | 1,864 (23.6%) | | 1,923 (24.3%) | 0.272 | 0.017 |
| Tenofovir alafenamide | 2,837 (28.3%) | 8,894 (2.1%) | **<0.001** | 0.785 | 1,437 (18.2%) | | 1,295 (16.4%) | **0.003** | 0.047 |
| Lamivudine | 1,494 (14.9%) | 1,593 (0.4%) | **<0.001** | 0.569 | 582 (7.4%) | | 564 (7.1%) | 0.581 | 0.009 |
| Entecavir | 255 (2.5%) | 14,177 (3.3%) | **<0.001** | 0.044 | 181 (2.3%) | | 336 (4.2%) | **0.001** | 0.091 |
| **NNRTIs** | 1,687 (16.8%) | - | - | - | 973 (12.3%) | | - | - | - |
| **PIs** | 2,024 (20.2%) | - | - | - | 1,081 (13.7%) | | - | - | - |
| **INSTIs** | 2,546 (25.4%) | - | - | - | 1,384 (17.5%) | | - | - | - |
| **HIV combinations** | 2,072 (20.6%) | - | - | - | 1,115 (14.1%) | | - | - | - |
| Laboratory markers |  |  |  |  |  | |  |  |  |
| CD4 count (cells/μL) |  |  |  |  |  | |  |  |  |
| Median (IQR) | 391 (482) | - | - | - | 389 (482) | | - | - | - |
| Available, n (%) | 4,244 (42.3%) | - | - | - | 2,969 (37.5%) | | - | - | - |
| CD4 percentage (%) |  |  |  |  |  | |  |  |  |
| Median (IQR) | 24 (19) | - | - | - | 24 (20) | | - | - | - |
| Available, n (%) | 4,680 (46.6%) | - | - | - | 3,270 (41.3%) | | - | - | - |
| HIV RNA (copies/mL) |  |  |  |  |  | |  |  |  |
| Median (IQR) | 1,761 (49,715) | - | - | - | 2,997 (61,413) | | - | - | - |
| Available, n (%) | 2,363 (23.5%) | - | - | - | 1,639 (20.7%) | | - | - | - |
| HBV DNA (IU/mL) |  |  |  |  |  | |  |  |  |
| Median (IQR) | 225 (6,486) | 208 (3,510) | **<0.001** |  | 230 (5,837) | | 514 (10,371) | **0.003** |  |
| Available, n (%) | 627 (6.2%) | 14,947 (3.5%) |  |  | 481 (6.1%) | | 564 (7.1%) |  |  |

Baseline demographic, clinical, mental health, and lifestyle-associated characteristics are shown before and after propensity score matching for adults with HIV/HBV coinfection and HBV monoinfection. Propensity score matching was performed using 1:1 nearest-neighbor matching without replacement. Data are presented as number (%) or mean ± standard deviation. P values are provided for descriptive purposes only. Standardized differences <0.10 indicate negligible imbalance between groups.

Laboratory availability is shown for the baseline period (≤6 months before index). Extending the lookback window would increase capture; thus, these percentages are minimum estimates and should not be interpreted as the true testing frequency in the cohorts.

Abbreviations: HBV, hepatitis B virus; HIV, human immunodeficiency virus; NRTI, nucleoside/nucleotide reverse transcriptase inhibitor; NNRTI, non‑nucleoside reverse transcriptase inhibitor; PI, protease inhibitor; INSTI, integrase strand transfer inhibitor; IQR, interquartile range.

**Supplementary Table S3.** Six-Month Lag Analysis for Incident Social Vulnerability Outcomes in Adults With HIV/HBV Coinfection Compared With HIV Monoinfection

| Outcomes | Cohorts | | | IR (95% CI)  (per 100 person-years) | | HR (95% CI) | p-Value |
| --- | --- | --- | --- | --- | --- | --- | --- |
|  | **Overall** | **HIV/HBV** | **HIV** | **HIV/HBV** | **HIV** |  |  |
| Primary outcome |  |  |  |  |  |  |  |
| Social vulnerability composite | 15,777 | 788 (10.2%) | 789 (9.8%) | 2.26 (2.11–2.42) | 1.90 (1.77–2.04) | 1.13 (1.03–1.25) | **0.013** |
| Material vulnerability |  |  |  |  |  |  |  |
| Material vulnerability composite | 16,631 | 421 (5.1%) | 397 (4.7%) | 1.12 (0.93–1.36) | 0.92 (0.76–1.12) | 1.21 (1.05–1.39) | **0.007** |
| Housing instability | 16,764 | 280 (3.4%) | 266 (3.2%) | 0.74 (0.59–0.93) | 0.61 (0.48–0.78) | 1.20 (1.01–1.42) | **0.037** |
| Food insecurity | 17,062 | 104 (1.2%) | 107 (1.3%) | 0.27 (0.19–0.39) | 0.24 (0.17–0.35) | 1.19 (0.91–1.56) | 0.206 |
| Transportation insecurity | 17,085 | 53 (0.6%) | 49 (0.6%) | 0.14 (0.08–0.23) | 0.11 (0.07–0.19) | 1.28 (0.86–1.89) | 0.219 |
| Financial insecurity | 17,036 | 142 (1.7%) | 108 (1.3%) | 0.37 (0.27–0.51) | 0.25 (0.17–0.36) | 1.59 (1.24–2.05) | **<0.001** |
| Employment insecurity | 16,993 | 99 (1.2%) | 86 (1.0%) | 0.26 (0.18–0.38) | 0.20 (0.13–0.30) | 1.27 (0.95–1.69) | 0.108 |
| Healthcare access  and engagement |  |  |  |  |  |  |  |
| Healthcare access composite | 16,267 | 516 (6.5%) | 471 (5.7%) | 1.43 (1.20–1.69) | 1.11 (0.93–1.32) | 1.21 (1.07–1.37) | **0.003** |
| Insurance instability | 17,075 | 51 (0.6%) | 27 (0.3%) | 0.13 (0.08–0.22) | 0.06 (0.03–0.13) | 2.22 (1.38–3.55) | **0.001** |
| Health access barriers | 17,077 | 40 (0.5%) | 33 (0.4%) | 0.10 (0.06–0.19) | 0.08 (0.04–0.15) | 1.39 (0.88–2.22) | 0.160 |
| Care disengagement or nonadherence | 16,651 | 388 (4.8%) | 376 (4.4%) | 1.05 (0.86–1.28) | 0.86 (0.70–1.05) | 1.20 (1.04–1.38) | **0.012** |
| Interpersonal adversity |  |  |  |  |  |  |  |
| Interpersonal adversity composite | 17,515 | 121 (1.4%) | 150 (1.7%) | 0.31 (0.22–0.43) | 0.33 (0.24–0.46) | 0.92 (0.72–1.17) | 0.500 |
| Adverse childhood experiences | 17,772 | 15 (0.2%) | 30 (0.3%) | 0.04 (0.01–0.10) | 0.07 (0.03–0.13) | 0.56 (0.30–1.03) | 0.060 |
| Primary support stressors | 17,669 | 86 (1.0%) | 112 (1.3%) | 0.22 (0.14–0.33) | 0.25 (0.17–0.35) | 0.89 (0.67–1.18) | 0.402 |
| Violence and victimization | 17,710 | 28 (0.3%) | 27 (0.3%) | 0.07 (0.03–0.14) | 0.06 (0.03–0.12) | 1.15 (0.68–1.96) | 0.602 |
| Social vulnerability |  |  |  |  |  |  |  |
| Social vulnerability composite | 17,467 | 183 (2.1%) | 192 (2.2%) | 0.46 (0.35–0.62) | 0.43 (0.32–0.56) | 1.17 (0.95–1.43) | 0.131 |
| Social isolation | 17,759 | 40 (0.5%) | 32 (0.4%) | 0.10 (0.06–0.18) | 0.07 (0.04–0.14) | 1.53 (0.96–2.44) | 0.074 |
| Social adjustment issues | 17,722 | 86 (1.0%) | 82 (0.9%) | 0.21 (0.14–0.33) | 0.18 (0.12–0.28) | 1.38 (1.01–1.87) | **0.040** |
| Legal and carceral issues | 17,748 | 38 (0.4%) | 49 (0.6%) | 0.09 (0.05–0.18) | 0.11 (0.06–0.18) | 0.95 (0.62–1.45) | 0.803 |

Incidence rates (IRs) and hazard ratios (HRs) are shown for incident social vulnerability outcomes among matched adults with HIV/HBV coinfection and HIV monoinfection after applying a 6-month post-index lag. Outcomes occurring within the first 6 months after index were excluded, and follow-up began after the lag period. Analyses excluded individuals with the corresponding outcome before the analytic time window. Hazard ratios were estimated using Cox proportional hazards models adjusted for age, sex, race or ethnicity, calendar year of index, and baseline medical comorbidities. IRs are reported per 100 person-years. The primary outcome was ≥1 incident social vulnerability indicator after the analytic start date.

Abbreviations: CI, confidence interval; HBV, hepatitis B virus; HIV, human immunodeficiency virus; HR, hazard ratio; IR, incidence rate.

**Supplementary Table S4.** Virologic and Treatment-Adjusted Analysis for Incident Social Vulnerability Outcomes in Adults With HIV/HBV Coinfection Compared With HIV Monoinfection

| Outcomes | Cohorts | | | IR (95% CI)  (per 100 person-years) | | HR (95% CI) | p-Value |
| --- | --- | --- | --- | --- | --- | --- | --- |
|  | **Overall** | **HIV/HBV** | **HIV** | **HIV/HBV** | **HIV** |  |  |
| Primary outcome |  |  |  |  |  |  |  |
| Social vulnerability composite | 16,412 | 660 (8.3%) | 621 (7.4%) | 1.83 (1.69–1.97) | 1.44 (1.33–1.56) | 1.29 (1.15–1.44) | **<0.001** |
| Material vulnerability |  |  |  |  |  |  |  |
| Material vulnerability composite | 17,613 | 375 (4.3%) | 301 (3.4%) | 0.95 (0.86–1.05) | 0.66 (0.59–0.74) | 1.50 (1.29–1.75) | **<0.001** |
| Housing instability | 17,830 | 237 (2.7%) | 180 (2.0%) | 0.59 (0.52–0.67) | 0.39 (0.34–0.45) | 1.57 (1.29–1.91) | **<0.001** |
| Food insecurity | 18,284 | 107 (1.2%) | 99 (1.1%) | 0.26 (0.21–0.31) | 0.21 (0.17–0.26) | 1.36 (1.03–1.79) | **0.029** |
| Transportation insecurity | 18,318 | 53 (0.6%) | 48 (0.5%) | 0.13 (0.10–0.17) | 0.10 (0.08–0.14) | 1.35 (0.91–2.00) | 0.136 |
| Financial insecurity | 18,245 | 138 (1.5%) | 96 (1.1%) | 0.34 (0.29–0.40) | 0.20 (0.17–0.25) | 1.80 (1.38–2.34) | **<0.001** |
| Employment insecurity | 18,195 | 87 (1.0%) | 72 (0.8%) | 0.21 (0.17–0.26) | 0.15 (0.12–0.19) | 1.40 (1.02–1.91) | **0.035** |
| Healthcare access  and engagement |  |  |  |  |  |  |  |
| Healthcare access composite | 17,160 | 433 (5.2%) | 400 (4.6%) | 1.14 (1.04–1.26) | 0.84 (0.76–0.93) | 1.27 (1.11–1.46) | **0.001** |
| Insurance instability | 18,337 | 53 (0.6%) | 21 (0.2%) | 0.13 (0.10–0.17) | 0.04 (0.03–0.07) | 2.98 (1.79–4.95) | **<0.001** |
| Health access barriers | 18,327 | 39 (0.4%) | 31 (0.3%) | 0.09 (0.07–0.13) | 0.07 (0.05–0.09) | 1.54 (0.96–2.48) | 0.074 |
| Care disengagement or nonadherence | 15,840 | 473 (6.1%) | 457 (5.7%) | 1.29 (1.18–1.42) | 1.06 (0.97–1.16) | 1.14 (1.00–1.30) | **0.043** |
| Interpersonal adversity |  |  |  |  |  |  |  |
| Interpersonal adversity composite | 16,379 | 135 (1.7%) | 155 (1.9%) | 0.36 (0.30–0.43) | 0.34 (0.29–0.40) | 0.96 (0.76–1.21) | 0.713 |
| Adverse childhood experiences | 16,555 | 18 (0.2%) | 25 (0.3%) | 0.05 (0.03–0.08) | 0.06 (0.04–0.08) | 0.79 (0.43–1.44) | 0.435 |
| Primary support stressors | 16,487 | 91 (1.1%) | 114 (1.4%) | 0.24 (0.20–0.30) | 0.27 (0.22–0.33) | 0.88 (0.67–1.16) | 0.367 |
| Violence and victimization | 16,502 | 35 (0.4%) | 34 (0.4%) | 0.09 (0.07–0.13) | 0.08 (0.06–0.11) | 1.11 (0.69–1.78) | 0.676 |
| Social vulnerability |  |  |  |  |  |  |  |
| Social vulnerability composite | 16,379 | 204 (2.5%) | 215 (2.6%) | 0.55 (0.48–0.63) | 0.51 (0.44–0.59) | 1.13 (0.93–1.37) | 0.218 |
| Social isolation | 16,552 | 42 (0.5%) | 39 (0.5%) | 0.11 (0.08–0.15) | 0.09 (0.07–0.13) | 1.25 (0.81–1.94) | 0.316 |
| Social adjustment issues | 16,530 | 96 (1.2%) | 90 (1.1%) | 0.26 (0.21–0.31) | 0.21 (0.17–0.26) | 1.35 (1.01–1.81) | **0.041** |
| Legal and carceral issues | 16,539 | 42 (0.5%) | 50 (0.6%) | 0.11 (0.08–0.15) | 0.12 (0.09–0.16) | 0.99 (0.66–1.50) | 0.971 |

Incidence rates (IRs) and hazard ratios (HRs) are shown for incident social vulnerability outcomes among matched adults with HIV/HBV coinfection and HIV monoinfection after additional adjustment for virologic markers and antiviral treatment. Analyses excluded individuals with the corresponding outcome before index. Hazard ratios were estimated using Cox proportional hazards models adjusted for age, sex, race or ethnicity, calendar year of index, baseline medical comorbidities, HIV RNA, CD4 count, HBV DNA, and antiretroviral therapy. IRs are reported per 100 person-years. The primary outcome was ≥1 incident social vulnerability indicator after index.

Abbreviations: CI, confidence interval; HBV, hepatitis B virus; HIV, human immunodeficiency virus; HR, hazard ratio; IR, incidence rate; IQR, interquartile range; RNA, ribonucleic acid. DNA, deoxyribonucleic acid.

**Supplementary Table S5.** Healthcare Utilization-Adjusted Analysis for Incident Social Vulnerability Outcomes in Adults With HIV/HBV Coinfection Compared With HIV Monoinfection

| Outcomes | Cohorts | | | IR (95% CI)  (per 100 person-years) | | HR (95% CI) | p-Value |
| --- | --- | --- | --- | --- | --- | --- | --- |
|  | **Overall** | **HIV/HBV** | **HIV** | **HIV/HBV** | **HIV** |  |  |
| Primary outcome |  |  |  |  |  |  |  |
| Social vulnerability composite | 18,100 | 1,360 (15.4%) | 1,255 (13.6%) | 33.6 (31.9–35.4) | 27.5 (26.0–29.1) | 1.21 (1.12–1.31) | **<0.001** |
| Material vulnerability |  |  |  |  |  |  |  |
| Material vulnerability composite | 19,704 | 520 (5.3%) | 494 (5.0%) | 11.6 (10.7–12.7) | 10.1 (9.3–11.1) | 1.15 (1.01–1.30) | **0.029** |
| Housing instability | 19,772 | 407 (4.1%) | 390 (3.9%) | 9.0 (8.2–9.9) | 8.0 (7.2–8.8) | 1.15 (1.00–1.32) | **0.050** |
| Food insecurity | 20,263 | 144 (1.4%) | 153 (1.5%) | 3.1 (2.6–3.7) | 3.1 (2.6–3.6) | 1.08 (0.86–1.36) | 0.495 |
| Transportation insecurity | 20,300 | 81 (0.8%) | 82 (0.8%) | 1.7 (1.4–2.2) | 1.6 (1.3–2.0) | 1.11 (0.82–1.51) | 0.501 |
| Financial insecurity | 20,126 | 276 (2.7%) | 211 (2.1%) | 6.0 (5.3–6.8) | 4.2 (3.7–4.9) | 1.45 (1.21–1.74) | **<0.001** |
| Employment insecurity | 20,172 | 114 (1.1%) | 120 (1.2%) | 2.5 (2.1–3.0) | 2.4 (2.0–2.9) | 1.03 (0.80–1.33) | 0.822 |
| Healthcare access  and engagement |  |  |  |  |  |  |  |
| Healthcare access composite | 19,108 | 721 (7.7%) | 685 (7.0%) | 17.0 (15.8–18.3) | 14.2 (13.2–15.4) | 1.15 (1.03–1.27) | **0.011** |
| Insurance instability | 20,298 | 63 (0.6%) | 33 (0.3%) | 1.4 (1.1–1.7) | 0.7 (0.5–0.9) | 2.08 (1.36–3.17) | **0.001** |
| Health access barriers | 20,296 | 59 (0.6%) | 62 (0.6%) | 1.3 (1.0–1.7) | 1.2 (0.9–1.6) | 1.03 (0.72–1.47) | 0.875 |
| Care disengagement or nonadherence | 19,220 | 658 (7.0%) | 628 (6.4%) | 15.2 (14.1–16.5) | 13.0 (12.0–14.1) | 1.13 (1.01–1.26) | **0.030** |
| Interpersonal adversity |  |  |  |  |  |  |  |
| Interpersonal adversity composite | 20,098 | 180 (1.8%) | 209 (2.1%) | 3.9 (3.4–4.5) | 4.2 (3.7–4.8) | 0.93 (0.76–1.14) | 0.476 |
| Adverse childhood experiences | 20,307 | 25 (0.2%) | 40 (0.4%) | 0.5 (0.4–0.8) | 0.8 (0.6–1.1) | 0.65 (0.40–1.07) | 0.091 |
| Primary support stressors | 20,232 | 123 (1.2%) | 142 (1.4%) | 2.7 (2.2–3.2) | 2.8 (2.4–3.4) | 0.95 (0.75–1.22) | 0.701 |
| Violence and victimization | 20,263 | 45 (0.4%) | 54 (0.5%) | 1.0 (0.7–1.3) | 1.1 (0.8–1.4) | 0.87 (0.59–1.30) | 0.501 |
| Social vulnerability |  |  |  |  |  |  |  |
| Social vulnerability composite | 20,065 | 233 (2.3%) | 238 (2.4%) | 5.1 (4.5–5.8) | 4.8 (4.2–5.5) | 1.12 (0.94–1.35) | 0.213 |
| Social isolation | 20,314 | 58 (0.6%) | 51 (0.5%) | 1.3 (1.0–1.6) | 1.0 (0.8–1.4) | 1.29 (0.88–1.88) | 0.194 |
| Social adjustment issues | 20,188 | 148 (1.5%) | 148 (1.5%) | 3.2 (2.7–3.8) | 3.0 (2.5–3.5) | 1.18 (0.93–1.48) | 0.167 |
| Legal and carceral issues | 20,294 | 51 (0.5%) | 75 (0.7%) | 1.1 (0.8–1.4) | 1.5 (1.2–1.9) | 0.75 (0.53–1.08) | 0.118 |

Incidence rates (IRs) and hazard ratios (HRs) are shown for incident social vulnerability outcomes among matched adults with HIV/HBV coinfection and HIV monoinfection after additional adjustment for virologic markers and antiviral treatment. Analyses excluded individuals with the corresponding outcome before index. Hazard ratios were estimated using Cox proportional hazards models adjusted for age, sex, race or ethnicity, calendar year of index, baseline medical comorbidities, HIV RNA, CD4 count, HBV DNA, and antiretroviral therapy. IRs are reported per 100 person-years. The primary outcome was ≥1 incident social vulnerability indicator after index.

Abbreviations: CI, confidence interval; HBV, hepatitis B virus; HIV, human immunodeficiency virus; HR, hazard ratio; IR, incidence rate; IQR, interquartile range; RNA, ribonucleic acid. DNA, deoxyribonucleic acid.

**Supplementary Table S6.** Six-Month Lag Analysis for Incident Social Vulnerability Outcomes in Adults With HIV/HBV Coinfection Compared With HBV Monoinfection

| Outcomes | Cohorts | | | IR (95% CI)  (per 100 person-years) | | HR (95% CI) | p-Value |
| --- | --- | --- | --- | --- | --- | --- | --- |
|  | **Overall** | **HIV/HBV** | **HIV** | **HIV/HBV** | **HIV** |  |  |
| Primary outcome |  |  |  |  |  |  |  |
| Social vulnerability composite | 17,110 | 748 (9.0%) | 443 (5.1%) | 1.95 (1.82–2.10) | 1.32 (1.20–1.45) | 1.50 (1.33–1.68) | **<0.001** |
| Material vulnerability |  |  |  |  |  |  |  |
| Material vulnerability composite | 18,476 | 410 (4.5%) | 191 (2.1%) | 0.99 (0.90–1.09) | 0.54 (0.47–0.62) | 1.80 (1.51–2.13) | **<0.001** |
| Housing instability | 18,709 | 262 (2.8%) | 101 (1.1%) | 0.63 (0.56–0.71) | 0.29 (0.24–0.35) | 2.17 (1.73–2.73) | **<0.001** |
| Food insecurity | 19,206 | 116 (1.2%) | 60 (0.6%) | 0.27 (0.23–0.32) | 0.17 (0.13–0.22) | 1.59 (1.16–2.17) | **0.003** |
| Transportation insecurity | 19,251 | 59 (0.6%) | 53 (0.6%) | 0.14 (0.11–0.18) | 0.15 (0.11–0.20) | 0.92 (0.63–1.33) | 0.659 |
| Financial insecurity | 19,140 | 144 (1.5%) | 80 (0.8%) | 0.34 (0.29–0.40) | 0.23 (0.18–0.28) | 1.48 (1.12–1.94) | **0.005** |
| Employment insecurity | 19,101 | 97 (1.0%) | 49 (0.5%) | 0.22 (0.18–0.27) | 0.14 (0.10–0.18) | 1.65 (1.17–2.33) | **0.004** |
| Healthcare access  and engagement |  |  |  |  |  |  |  |
| Healthcare access composite | 17,903 | 505 (5.8%) | 288 (3.2%) | 1.26 (1.15–1.38) | 0.82 (0.73–0.92) | 1.55 (1.34–1.79) | **<0.001** |
| Insurance instability | 19,233 | 54 (0.6%) | 27 (0.3%) | 0.12 (0.09–0.16) | 0.08 (0.05–0.11) | 1.64 (1.03–2.61) | **0.034** |
| Health access barriers | 19,241 | 44 (0.5%) | 34 (0.4%) | 0.10 (0.07–0.14) | 0.10 (0.07–0.13) | 1.08 (0.69–1.68) | 0.748 |
| Care disengagement or nonadherence | 18,029 | 456 (5.2%) | 239 (2.6%) | 1.12 (1.02–1.23) | 0.67 (0.59–0.76) | 1.71 (1.46–2.00) | **<0.001** |
| Interpersonal adversity |  |  |  |  |  |  |  |
| Interpersonal adversity composite | 18,982 | 144 (1.5%) | 84 (0.9%) | 0.34 (0.29–0.40) | 0.24 (0.19–0.30) | 1.46 (1.11–1.91) | **0.006** |
| Adverse childhood experiences | 19,252 | 18 (0.2%) | 17 (0.2%) | 0.04 (0.02–0.07) | 0.05 (0.03–0.08) | 0.91 (0.47–1.76) | 0.767 |
| Primary support stressors | 19,120 | 105 (1.1%) | 64 (0.7%) | 0.24 (0.20–0.29) | 0.18 (0.14–0.23) | 1.39 (1.02–1.89) | **0.038** |
| Violence and victimization | 19,218 | 32 (0.3%) | 13 (0.1%) | 0.07 (0.05–0.11) | 0.04 (0.02–0.06) | 2.10 (1.10–3.99) | **0.021** |
| Social vulnerability |  |  |  |  |  |  |  |
| Social vulnerability composite | 18,910 | 192 (2.0%) | 151 (1.6%) | 0.45 (0.39–0.52) | 0.42 (0.36–0.49) | 1.07 (0.86–1.32) | 0.542 |
| Social isolation | 19,250 | 46 (0.5%) | 37 (0.4%) | 0.11 (0.08–0.14) | 0.10 (0.07–0.14) | 1.04 (0.67–1.60) | 0.861 |
| Social adjustment issues | 19,190 | 88 (0.9%) | 58 (0.6%) | 0.20 (0.16–0.25) | 0.16 (0.12–0.21) | 1.26 (0.91–1.76) | 0.167 |
| Legal and carceral issues | 19,227 | 39 (0.4%) | 29 (0.3%) | 0.09 (0.06–0.12) | 0.08 (0.06–0.12) | 1.11 (0.68–1.79) | 0.679 |

Incidence rates (IRs) and hazard ratios (HRs) are shown for incident social vulnerability outcomes among matched adults with HIV/HBV coinfection and HBV monoinfection after applying a 6-month post-index lag. Outcomes occurring within the first 6 months after index were excluded, and follow-up began after the lag period. Analyses excluded individuals with the corresponding outcome before the analytic time window. Hazard ratios were estimated using Cox proportional hazards models adjusted for age, sex, race or ethnicity, calendar year of index, and baseline medical comorbidities. IRs are reported per 100 person-years. The primary outcome was ≥1 incident social vulnerability indicator after the analytic start date.

Abbreviations: CI, confidence interval; HBV, hepatitis B virus; HIV, human immunodeficiency virus; HR, hazard ratio; IR, incidence rate.

**Supplementary Table S7.** Virologic and Treatment-Adjusted Analysis for Incident Social Vulnerability Outcomes in Adults With HIV/HBV Coinfection Compared With HBV Monoinfection

| Outcomes | Cohorts | | | IR (95% CI)  (per 100 person-years) | | HR (95% CI) | p-Value |
| --- | --- | --- | --- | --- | --- | --- | --- |
|  | **Overall** | **HIV/HBV** | **HIV** | **HIV/HBV** | **HIV** |  |  |
| Primary outcome |  |  |  |  |  |  |  |
| Social vulnerability composite | 14,591 | 743 (10.3%) | 431 (5.8%) | 2.3 (2.1–2.4) | 1.5 (1.4–1.7) | 1.61 (1.43–1.81) | **<0.001** |
| Material vulnerability |  |  |  |  |  |  |  |
| Material vulnerability composite | 15,436 | 380 (4.9%) | 180 (2.3%) | 1.1 (1.0–1.2) | 0.6 (0.5–0.7) | 1.89 (1.58–2.26) | **<0.001** |
| Housing instability | 15,557 | 257 (3.3%) | 109 (1.4%) | 0.7 (0.6–0.8) | 0.4 (0.3–0.5) | 2.10 (1.68–2.63) | **<0.001** |
| Food insecurity | 15,830 | 88 (1.1%) | 38 (0.5%) | 0.3 (0.2–0.3) | 0.1 (0.1–0.2) | 2.01 (1.38–2.95) | **<0.001** |
| Transportation insecurity | 15,854 | 45 (0.6%) | 30 (0.4%) | 0.1 (0.1–0.2) | 0.1 (0.1–0.1) | 1.32 (0.83–2.09) | 0.239 |
| Financial insecurity | 15,806 | 123 (1.6%) | 66 (0.8%) | 0.4 (0.3–0.4) | 0.2 (0.2–0.3) | 1.64 (1.22–2.22) | **0.001** |
| Employment insecurity | 15,770 | 92 (1.2%) | 47 (0.6%) | 0.3 (0.2–0.3) | 0.2 (0.1–0.2) | 1.72 (1.21–2.44) | **0.002** |
| Healthcare access  and engagement |  |  |  |  |  |  |  |
| Healthcare access composite | 15,113 | 500 (6.7%) | 268 (3.5%) | 1.5 (1.3–1.6) | 0.9 (0.8–1.0) | 1.72 (1.49–2.00) | **<0.001** |
| Insurance instability | 15,836 | 49 (0.6%) | 26 (0.3%) | 0.1 (0.1–0.2) | 0.1 (0.1–0.1) | 1.63 (1.02–2.63) | **0.041** |
| Health access barriers | 15,848 | 36 (0.5%) | 29 (0.4%) | 0.1 (0.1–0.1) | 0.1 (0.1–0.1) | 1.09 (0.67–1.78) | 0.727 |
| Care disengagement or nonadherence | 15,521 | 475 (6.2%) | 240 (3.0%) | 1.4 (1.2–1.5) | 0.8 (0.7–0.9) | 1.86 (1.59–2.17) | **<0.001** |
| Interpersonal adversity |  |  |  |  |  |  |  |
| Interpersonal adversity composite | 16,006 | 141 (1.8%) | 89 (1.1%) | 0.4 (0.3–0.4) | 0.3 (0.2–0.3) | 1.41 (1.08–1.84) | **0.010** |
| Adverse childhood experiences | 16,212 | 18 (0.2%) | 13 (0.2%) | 0.0 (0.0–0.1) | 0.0 (0.0–0.1) | 1.24 (0.61–2.54) | 0.550 |
| Primary support stressors | 16,150 | 96 (1.2%) | 62 (0.8%) | 0.3 (0.2–0.3) | 0.2 (0.2–0.3) | 1.38 (1.00–1.90) | **0.049** |
| Violence and victimization | 16,137 | 35 (0.4%) | 25 (0.3%) | 0.1 (0.1–0.1) | 0.1 (0.1–0.1) | 1.26 (0.75–2.10) | 0.382 |
| Social vulnerability |  |  |  |  |  |  |  |
| Social vulnerability composite | 16,024 | 176 (2.2%) | 155 (1.9%) | 0.5 (0.4–0.6) | 0.5 (0.4–0.6) | 1.02 (0.82–1.27) | 0.864 |
| Social isolation | 16,226 | 44 (0.5%) | 27 (0.3%) | 0.1 (0.1–0.1) | 0.1 (0.1–0.1) | 1.45 (0.90–2.34) | 0.126 |
| Social adjustment issues | 16,200 | 76 (0.9%) | 65 (0.8%) | 0.2 (0.2–0.3) | 0.2 (0.2–0.3) | 1.05 (0.76–1.47) | 0.761 |
| Legal and carceral issues | 16,200 | 32 (0.4%) | 19 (0.2%) | 0.1 (0.1–0.1) | 0.1 (0.0–0.1) | 1.48 (0.84–2.61) | 0.174 |

Incidence rates (IRs) and hazard ratios (HRs) are shown for incident social vulnerability outcomes among matched adults with HIV/HBV coinfection and HBV monoinfection after additional adjustment for virologic markers and antiviral treatment. Analyses excluded individuals with the corresponding outcome before index. Hazard ratios were estimated using Cox proportional hazards models adjusted for age, sex, race or ethnicity, calendar year of index, baseline medical comorbidities, HIV RNA, CD4 count, HBV DNA, and antiretroviral therapy. IRs are reported per 100 person-years. The primary outcome was ≥1 incident social vulnerability indicator after index.

Abbreviations: CI, confidence interval; HBV, hepatitis B virus; HIV, human immunodeficiency virus; HR, hazard ratio; IR, incidence rate; IQR, interquartile range; RNA, ribonucleic acid; DNA, deoxyribonucleic acid.

**Supplementary Table S8.** Healthcare Utilization-Adjusted Analysis for Incident Social Vulnerability Outcomes in Adults With HIV/HBV Coinfection Compared With HBV Monoinfection

| Outcomes | Cohorts | | | IR (95% CI)  (per 100 person-years) | | HR (95% CI) | p-Value |
| --- | --- | --- | --- | --- | --- | --- | --- |
|  | **Overall** | **HIV/HBV** | **HIV** | **HIV/HBV** | **HIV** |  |  |
| Primary outcome |  |  |  |  |  |  |  |
| Social vulnerability composite | 18,413 | 1,013 (11.2%) | 534 (5.7%) | 24.5 (23.1–26.1) | 14.6 (13.4–15.9) | 1.75 (1.58–1.95) | **<0.001** |
| Material vulnerability |  |  |  |  |  |  |  |
| Material vulnerability composite | 19,675 | 521 (5.3%) | 253 (2.6%) | 11.6 (10.7–12.7) | 6.5 (5.8–7.4) | 1.81 (1.56–2.10) | **<0.001** |
| Housing instability | 19,843 | 342 (3.5%) | 154 (1.5%) | 7.6 (6.8–8.4) | 3.9 (3.4–4.6) | 1.95 (1.61–2.36) | **<0.001** |
| Food insecurity | 20,286 | 144 (1.4%) | 80 (0.8%) | 3.1 (2.6–3.7) | 2.0 (1.6–2.5) | 1.55 (1.18–2.03) | **0.002** |
| Transportation insecurity | 20,320 | 81 (0.8%) | 57 (0.6%) | 1.7 (1.4–2.2) | 1.4 (1.1–1.9) | 1.23 (0.88–1.73) | 0.224 |
| Financial insecurity | 20,245 | 172 (1.7%) | 96 (0.9%) | 3.7 (3.2–4.3) | 2.4 (2.0–2.9) | 1.54 (1.20–1.98) | **0.001** |
| Employment insecurity | 20,200 | 115 (1.1%) | 55 (0.5%) | 2.5 (2.1–3.0) | 1.4 (1.0–1.8) | 1.83 (1.32–2.52) | **<0.001** |
| Healthcare access  and engagement |  |  |  |  |  |  |  |
| Healthcare access composite | 19,112 | 718 (7.6%) | 357 (3.7%) | 16.8 (15.6–18.1) | 9.4 (8.5–10.4) | 1.84 (1.62–2.09) | **<0.001** |
| Insurance instability | 20,299 | 63 (0.6%) | 28 (0.3%) | 1.4 (1.1–1.7) | 0.7 (0.5–1.0) | 1.93 (1.24–3.01) | **0.003** |
| Health access barriers | 20,289 | 59 (0.6%) | 52 (0.5%) | 1.3 (1.0–1.6) | 1.3 (1.0–1.7) | 0.98 (0.68–1.43) | 0.924 |
| Care disengagement or nonadherence | 19,231 | 657 (7.0%) | 328 (3.4%) | 15.2 (14.1–16.4) | 8.6 (7.7–9.6) | 1.84 (1.61–2.10) | **<0.001** |
| Interpersonal adversity |  |  |  |  |  |  |  |
| Interpersonal adversity composite | 20,083 | 180 (1.8%) | 93 (0.9%) | 3.9 (3.4–4.5) | 2.4 (1.9–2.9) | 1.69 (1.32–2.17) | **<0.001** |
| Adverse childhood experiences | 20,303 | 25 (0.2%) | 16 (0.2%) | 0.5 (0.4–0.8) | 0.4 (0.2–0.7) | 1.34 (0.72–2.52) | 0.357 |
| Primary support stressors | 20,205 | 123 (1.2%) | 74 (0.7%) | 2.7 (2.2–3.2) | 1.9 (1.5–2.4) | 1.44 (1.08–1.93) | **0.012** |
| Violence and victimization | 20,270 | 43 (0.4%) | 15 (0.1%) | 0.9 (0.7–1.3) | 0.4 (0.2–0.6) | 2.52 (1.40–4.54) | **0.001** |
| Social vulnerability |  |  |  |  |  |  |  |
| Social vulnerability composite | 20,032 | 232 (2.3%) | 174 (1.7%) | 5.1 (4.5–5.8) | 4.4 (3.8–5.1) | 1.15 (0.95–1.41) | 0.153 |
| Social isolation | 20,302 | 58 (0.6%) | 36 (0.4%) | 1.3 (1.0–1.6) | 0.9 (0.6–1.3) | 1.38 (0.91–2.09) | 0.126 |
| Social adjustment issues | 20,267 | 100 (1.0%) | 75 (0.7%) | 2.2 (1.8–2.6) | 1.9 (1.5–2.4) | 1.14 (0.85–1.54) | 0.383 |
| Legal and carceral issues | 20,282 | 51 (0.5%) | 36 (0.4%) | 1.1 (0.8–1.4) | 0.9 (0.6–1.3) | 1.21 (0.79–1.85) | 0.391 |

Incidence rates (IRs) and hazard ratios (HRs) are shown for incident social vulnerability outcomes among matched adults with HIV/HBV coinfection and HBV monoinfection after additional adjustment for virologic markers and antiviral treatment. Analyses excluded individuals with the corresponding outcome before index. Hazard ratios were estimated using Cox proportional hazards models adjusted for age, sex, race or ethnicity, calendar year of index, baseline medical comorbidities, HIV RNA, CD4 count, HBV DNA, and antiretroviral therapy. IRs are reported per 100 person-years. The primary outcome was ≥1 incident social vulnerability indicator after index.

Abbreviations: CI, confidence interval; HBV, hepatitis B virus; HIV, human immunodeficiency virus; HR, hazard ratio; IR, incidence rate; IQR, interquartile range; RNA, ribonucleic acid; DNA, deoxyribonucleic acid.
