## Supplementary File for "Social Determinants of Health in HIV/HBV Coinfection Compared with HIV and HBV Monoinfection: A Framework for Dynamic Individual-Level Social Vulnerability"

Supplementary File 1. Cohort Definitions, Baseline Variables, and Outcome Code Lists

Study: Incident Social Determinants of Health in HIV/HBV Coinfection Compared With HIV and HBV Monoinfection: A Framework for Dynamic Individual-Level Social Vulnerability

**Document Overview**

This supplement provides boxed definitions and code lists for cohort creation, baseline variables used for propensity score matching, incident social vulnerability outcomes, and prespecified sensitivity analyses. Outcomes were assessed after index, and individuals with the corresponding outcome documented before follow-up were excluded from each outcome-specific analysis.

### 1. Data Source and Analytic Framework

**Data Source and Time Window**

Data source: deidentified multi-institutional electronic health record network (TriNetX Research Network).

Eligible population: adults aged 18 years or older with qualifying cohort-defining events recorded on or after January 1, 2010.

Analytic comparisons: HIV/HBV coinfection vs HIV monoinfection; HIV/HBV coinfection vs HBV monoinfection.

Follow-up: began 1 day after the index date and continued until incident outcome occurrence or last available EHR activity.

### 2. Cohort Creation

**Cohort 1: HIV/HBV Coinfection**

Adults aged 18 years or older with evidence of both HIV and chronic HBV on or after January 1, 2010.

Inclusion: at least one HIV diagnosis code and at least one chronic HBV diagnosis code or HBV laboratory criterion.

Exclusions: HCV infection or positive HCV RNA, liver transplantation, or transplant-related complications.

**Cohort 2: HIV Monoinfection**

Adults aged 18 years or older with evidence of HIV on or after January 1, 2010 and no evidence of chronic HBV.

Inclusion: at least one HIV diagnosis code.

Exclusions: HBV diagnosis/laboratory evidence, HCV infection or positive HCV RNA, liver transplantation, or transplant-related complications.

**Cohort 3: HBV Monoinfection**

Adults aged 18 years or older with evidence of chronic HBV on or after January 1, 2010 and no evidence of HIV.

Inclusion: at least one chronic HBV diagnosis code or HBV laboratory criterion.

Exclusions: HIV diagnosis codes, HCV infection or positive HCV RNA, liver transplantation, or transplant-related complications.

Table S1. Cohort-defining and exclusion code lists

| **Construct** | **Code system** | **Codes** | **Definition / notes** |
| --- | --- | --- | --- |
| HIV infection | ICD-10-CM diagnoses | B20; Z21; O98.719; O98.72; B97.35 | HIV disease, asymptomatic HIV infection status, HIV complicating pregnancy/childbirth, or HIV-2-related disease. |
| Chronic HBV infection | ICD-10-CM diagnoses | B18.0; B18.1 | Chronic viral hepatitis B with or without delta-agent. |
| Chronic HBV infection | LOINC laboratory criteria | 5195-3; 58452-4; 95234-1; 5196-1; 47364-5; 11258-1; 95236-6; 5007-0; 5009-6; 29610-3; 20442-0; 49360-1; 49366-8; 49600-0; 42595-9; 48398-2; 45161-7 | Positive HBsAg or detectable HBV DNA above assay threshold. |
| HCV exclusion | ICD-10-CM diagnosis | B18.2 | Chronic viral hepatitis C. |
| HCV exclusion | LOINC laboratory criteria | 5012-0; 5010-4; 48576-3; 11259-9; 49376-7; 49379-1; 49380-9; 10676-5; 49375-9; 50023-1; 49372-6; 49605-9; 11011-4; 20416-4; 29609-5; 38180-6; 42617-1; 49758-6; 47252-2 | Positive or detectable HCV RNA. |
| Liver transplant exclusion | ICD-10-CM diagnoses | Z94.4; Z48.23; T86.4; T86.40; T86.41; T86.42; T86.43; T86.49 | Liver transplant status, aftercare, rejection, failure, infection, unspecified complication, or other transplant complications. |

Table S2. Index date, follow-up, and outcome ascertainment

| **Element** | **Definition** |
| --- | --- |
| Index date | First date on which an individual met qualifying cohort criteria. For HIV/HBV coinfection, this was the first date on which criteria for both HIV and chronic HBV were met. For monoinfection cohorts, this was the first qualifying HIV or HBV event. |
| Follow-up start | One day after the index date. |
| Outcome ascertainment | Outcomes occurring after the start of follow-up were included. For each outcome, individuals with the same outcome documented before follow-up were excluded from that outcome-specific analysis. |
| Censoring | Follow-up continued until first occurrence of the outcome of interest or last available EHR activity. |

### 3. Baseline Variables Used for Propensity Score Matching

**Propensity Score Matching**

Propensity score matching was performed separately for each comparison using 1:1 nearest-neighbor matching without replacement. Variables were selected a priori and grouped into demographics, baseline medical comorbidities, baseline mental health diagnoses, and baseline substance use-related conditions.

Table S3. Baseline variables included in propensity score matching

| **Category** | **Variables** |
| --- | --- |
| Demographics | Age at index; male; female; Hispanic or Latino; unknown ethnicity; Black or African American; White; unknown race; Asian; other race |
| Baseline medical comorbidities | Diseases of liver; diabetes mellitus; hypertensive diseases; heart failure; ischemic heart diseases; cerebrovascular diseases; chronic lower respiratory diseases; chronic kidney disease; neoplasms |
| Baseline mental health diagnoses | Major depressive disorder; bipolar disorder; anxiety disorders; post-traumatic stress disorder; schizophrenia, schizotypal, delusional, and other non-mood psychotic disorders |
| Baseline substance use-related conditions | Nicotine dependence; alcohol-related disorders; opioid-related disorders; cocaine-related disorders; cannabis-related disorders; other psychoactive substance-related disorders; other stimulant-related disorders |

Table S4. Baseline condition code groups

| **Variable** | **Code system** | **Codes** |
| --- | --- | --- |
| Hypertensive diseases | ICD-10-CM | I10-I11 |
| Ischemic heart diseases | ICD-10-CM | I20-I25 |
| Heart failure | ICD-10-CM | I50 |
| Diabetes mellitus | ICD-10-CM | E08-E13 |
| Chronic lower respiratory diseases | ICD-10-CM | J40-J4A |
| Chronic kidney disease | ICD-10-CM | N18 |
| Diseases of liver | ICD-10-CM | K70-K77 |
| Neoplasms | ICD-10-CM | C00-D49 |
| Nicotine dependence | ICD-10-CM | F17 |
| Alcohol-related disorders | ICD-10-CM | F10 |
| Opioid-related disorders | ICD-10-CM | F11 |
| Cannabis-related disorders | ICD-10-CM | F12 |
| Cocaine-related disorders | ICD-10-CM | F14 |
| Other stimulant-related disorders | ICD-10-CM | F15 |
| Other psychoactive substance-related disorders | ICD-10-CM | F19 |
| Major depressive disorder | ICD-10-CM | F32-F33 |
| Anxiety disorders | ICD-10-CM | F40-F41 |
| Post-traumatic stress disorder and trauma-related disorders | ICD-10-CM | F43 |
| Bipolar disorder | ICD-10-CM | F31 |
| Schizophrenia and other psychotic disorders | ICD-10-CM | F20-F29 |

### 4. Outcome Definitions and Code Lists

**Primary Outcome**

The primary outcome was an incident social vulnerability composite, defined as the first documentation of any qualifying SDoH indicator from any prespecified domain after the index date.

**Outcome Domains**

Secondary outcomes included four domain-specific composites and individual component outcomes. Domain-specific composites were defined as the first documentation of any component outcome within the domain after index. Individual component outcomes were analyzed separately.

**Material Vulnerability**

Unmet basic resource needs that may constrain stable living conditions, nutrition, transportation, employment, and the ability to sustain longitudinal HIV/HBV care. Components: housing instability, food insecurity, transportation insecurity, financial insecurity, and employment insecurity.

**Healthcare Access and Engagement**

Barriers to obtaining, financing, and remaining engaged in medical care. Components: insurance instability, healthcare access barriers, and care disengagement or nonadherence.

**Interpersonal Adversity**

Relational and family-level stressors that may affect disclosure, support, safety, and self-management. Components: adverse childhood experiences, primary support stressors, and violence or victimization.

**Social Vulnerability**

Broader social-contextual problems distinct from direct interpersonal relationships, including disruptions in social integration, community participation, adaptation to social environments, and legal or carceral circumstances. Components: social isolation, social adjustment issues, and legal or carceral issues.

Table S5. Outcome definitions and code lists - Primary outcome

| **Outcome** | **Operational definition** | **Codes** |
| --- | --- | --- |
| Social vulnerability composite | Any qualifying indicator from material vulnerability, healthcare access and engagement, interpersonal adversity, or social vulnerability domains. | ICD-10-CM: Z56; Z59; Z60; Z62; Z63; Z65; Z75; Z91.1; Z91.A; T74; T76; Z04.4; Z04.7 \| ICD-9-CM: V60; V60.8; V60.89 |

Table S5.1. Material vulnerability outcome definitions and code lists

| **Outcome** | **Operational definition** | **Codes** |
| --- | --- | --- |
| Material vulnerability composite | Any material vulnerability component. | ICD-10-CM: Z56; Z59.0; Z59.1; Z59.2; Z59.3; Z59.4; Z59.5; Z59.6; Z59.8; Z59.9; Z59.72; Z59.81; Z59.82; Z59.86; Z59.87; Z59.89 \| ICD-9-CM: V60; V60.8; V60.89 |
| Housing instability | Homelessness, inadequate housing, housing instability, discord with neighbors/landlord, residential institution issues, or unspecified housing/economic circumstances. | ICD-10-CM: Z59.0; Z59.1; Z59.2; Z59.3; Z59.9; Z59.81; Z59.89 \| ICD-9-CM: V60; V60.8; V60.89 |
| Food insecurity | Lack of adequate food. | ICD-10-CM: Z59.4 |
| Transportation insecurity | Transportation insecurity. | ICD-10-CM: Z59.82 |
| Financial insecurity | Financial insecurity, material hardship, insufficient welfare support, extreme poverty, low income, or related housing/economic hardship. | ICD-10-CM: Z59.86; Z59.87; Z59.72; Z59.5; Z59.6; Z59.89 \| ICD-9-CM: V60; V60.8; V60.89 |
| Employment insecurity | Problems related to employment and unemployment. | ICD-10-CM: Z56 |

Table S5.2. Healthcare access and engagement outcome definitions and code lists

| **Outcome** | **Operational definition** | **Codes** |
| --- | --- | --- |
| Healthcare access composite | Any healthcare access and engagement component. | ICD-10-CM: Z75; Z91.1; Z91.A; Z59.71 |
| Insurance instability | Insufficient health insurance coverage. | ICD-10-CM: Z59.71 |
| Healthcare access barriers | Problems related to medical facilities and other health care. | ICD-10-CM: Z75 |
| Care disengagement or nonadherence | Patient or caregiver noncompliance with medical treatment and regimen. | ICD-10-CM: Z91.1; Z91.A |

Table S5.3. Interpersonal adversity outcome definitions and code lists

| **Outcome** | **Operational definition** | **Codes** |
| --- | --- | --- |
| Interpersonal adversity composite | Any interpersonal adversity component. | ICD-10-CM: Z62; Z63; T74; T76; Z04.4; Z04.7 |
| Adverse childhood experiences | Problems related to upbringing. | ICD-10-CM: Z62 |
| Primary support stressors | Problems related to primary support group, including family circumstances. | ICD-10-CM: Z63 |
| Violence or victimization | Confirmed or suspected abuse/neglect/maltreatment or encounters following alleged rape or physical abuse. | ICD-10-CM: T74; T76; Z04.4; Z04.7 |

Table S5.4. Social vulnerability outcome definitions and code lists

| **Outcome** | **Operational definition** | **Codes** |
| --- | --- | --- |
| Social vulnerability composite | Any social vulnerability domain component. | ICD-10-CM: Z60; Z65 |
| Social isolation | Living alone, social exclusion/rejection, or target of perceived discrimination/persecution. | ICD-10-CM: Z60.2; Z60.4; Z60.5 |
| Social adjustment issues | Adjustment to life-cycle transitions, acculturation difficulty, other social environment problems, or unspecified social environment problem. | ICD-10-CM: Z60.0; Z60.3; Z60.8; Z60.9 |
| Legal and carceral issues | Civil/criminal proceedings, imprisonment/incarceration, release from prison, or other legal circumstances. | ICD-10-CM: Z65.0; Z65.1; Z65.2; Z65.3 |

### 5. Analysis Settings and Sensitivity Analyses

Table S6. Analysis settings and sensitivity analyses

| **Analysis element** | **Specification** |
| --- | --- |
| Analysis type | Compare outcomes using risk and time-to-event analyses. Hazard ratios were estimated using Cox proportional hazards models. |
| Prevalent outcome exclusion | Analyses excluded individuals with the corresponding outcome documented before the outcome ascertainment window. |
| Incidence rates | Reported per 100 person-years with 95% confidence intervals. |
| Primary sensitivity analysis | 6-month landmark analysis restricted outcome ascertainment to events occurring at least 6 months after index. |
| Virologic/treatment-adjusted sensitivity analysis | Models additionally adjusted for baseline virologic markers and treatment variables where applicable. |

Table S7. Additional variables used in virologic and treatment-adjusted sensitivity analyses

| **Construct** | **Variables / medications** |
| --- | --- |
| HIV disease markers | CD4 count category; HIV RNA suppression status |
| HIV treatment | Antiretroviral therapy use |
| HBV disease markers | HBV DNA detectability |
| HBV-active antiviral therapy | Tenofovir disoproxil fumarate; tenofovir alafenamide; entecavir; lamivudine |

### Abbreviations

**Abbreviations**

CI, confidence interval; EHR, electronic health record; HBV, hepatitis B virus; HCV, hepatitis C virus; HCC, hepatocellular carcinoma; HIV, human immunodeficiency virus; ICD-9-CM, International Classification of Diseases, Ninth Revision, Clinical Modification; ICD-10-CM, International Classification of Diseases, Tenth Revision, Clinical Modification; LOINC, Logical Observation Identifiers Names and Codes; SDoH, social determinants of health.
